# Higher omega-3 DHA-plasmalogen phospholipid levels across early life mediate the benefits of maternal healthy diet and breastfeeding on infant neurocognition

**DOI:** 10.64898/2026.03.22.26348566

**Authors:** Kristina Vacy, Katherine Drummond, Sarah Thomson, Andrea Gogos, Christos Symeonides, David Burgner, Toby Mansell, Richard Saffery, Mimi LK Tang, Martin O’Hely, Alexandra D. George, Sudip Paul, Peter J Meikle, Wah Chin Boon, Satvika Burugupalli, Peter Vuillermin, Anne-Louise Ponsonby, the Barwon Infant Study Investigator Group

**Author notes:** These authors contributed equally (Peter Vuillermin, Anne-Louise Ponsonby).

## Abstract

Early-life lipid profiles may influence neurodevelopment, yet the underlying biochemical pathways are not well defined. Using serial lipidomic data from 1,074 mother–child pairs in the Barwon Infant Study, we examined associations between maternal and infant lipid networks and cognitive and language outcomes at two years. A shared lipid network enriched in DHA-carrying phosphatidylethanolamine plasmalogens (PEP-DHA) was associated with higher subsequent cognition and language at age two years. These findings were distinct from total DHA levels alone. Antenatally, maternal PEP-DHA partly mediated the relationship between prenatal diet quality and cognition and language. Postnatally, the beneficial effect of breastfeeding on neurocognition was substantially mediated by PEP-DHA levels in infant plasma at 6 months, while infant PEP-DHA mediated the effect of breastfeeding on receptive language. These findings suggest that DHA-containing plasmalogens may represent key lipid mediators linking early nutrition with optimal neurodevelopment.

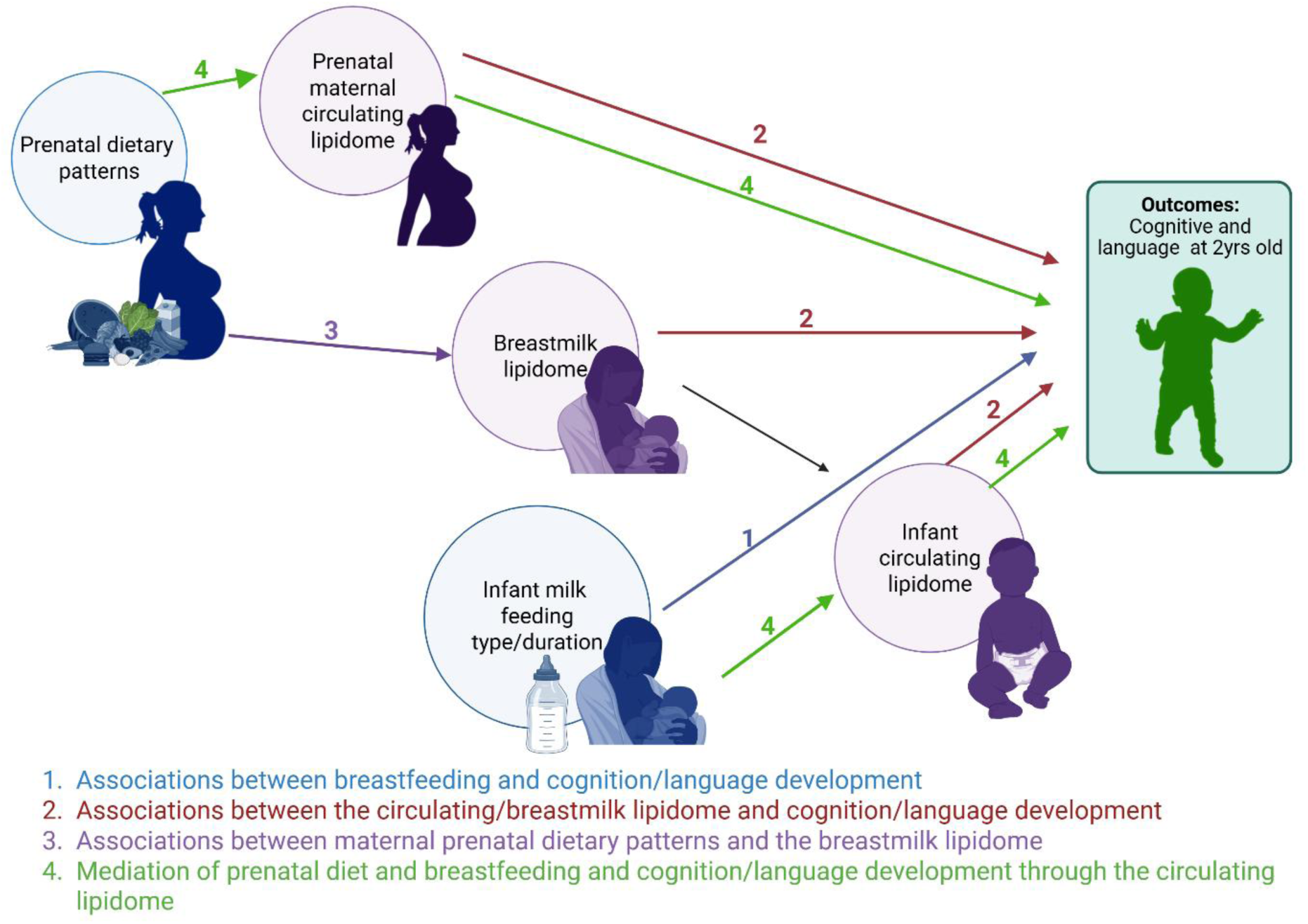

## Introduction

Optimal nutrition during early life is increasingly recognised as a critical determinant of healthy neurodevelopment. Maternal diet during pregnancy and the nutritional composition of human breast milk provide essential lipid nutrients that may support the developing brain^1, 2^. Lipids constitute approximately 50-60% of the brain’s dry weight^3, 4^ underscoring their structural and functional importance in neural tissue. Metabolomics offers a powerful approach to explore how these nutrients influence neurodevelopment, with lipidomics—focused on the comprehensive profiling of lipid species—representing a key domain within this field. The role of the omega-3 fatty acid docosahexaenoic acid (DHA; 22:6n-3) is of particular interest as it accumulates specifically in the membrane lipids of the brain and retina in early life, where it is important to neural and visual function^5^. The period of most rapid DHA accretion occurs from the third trimester through the first six months of postnatal life, coinciding with breastfeeding^6, 7^. As DHA can be synthesised only from the dietary omega-3 precursor α-linolenic acid (ALA; 18:3n-3), its availability depends heavily on maternal intake and dietary sources such as fish oil, which also provide eicosapentaenoic acid (EPA; 20:5n-3).

Beyond DHA itself, lipid classes in which DHA is incorporated may are also biologically significant^8^. One such example is phosphatidylethanolamine plasmalogen (PE(P)), a vinyl ether-linked phospholipid implicated in the developmental accumulation of DHA in the brain^8, 9^. These are biologically active in its own right and also serves as an important carrier of DHA and other omega-3s^10^. Ether lipids are characterised by an ether linkage on their glycerol backbone unlike the more common ester bond found in other lipids. Plasmalogens are a subclass of ether lipids that contain a vinyl ether bond (**Extended Online Figure 1**). Phosphatidylethanolamine plasmalogens (PE(P); also termed alkenyl-PE or plasmenyl-PE; also abbreviated PlsEtn), are the second-most abundant ether lipid class in breast milk, following the ether lipid class alkyl-diacylglycerols (TG(O))^11^. PE(P) species are also highly enriched in neural tissue^12^, comprising approximately 60% of ethanolamine phospholipids in grey matter and up to 80% in white matter/myelin^13^. DHA within PE(P) species (“PEP-DHA”) features a PE headgroup, a vinyl ether bond with a saturated or monounsaturated fatty acid at the stereospecific numbering 1 (sn-1) position, and DHA (22:6) fatty acid at the sn-2 position bond.

The World Health Organization (WHO) recommends exclusive breastfeeding for the first six months and promotes continued breastfeeding up to two years^14, 15^ reflecting strong evidence for its neurodevelopmental and health benefits^1, 15, 16, 17, 18, 19, 20, 21, 22, 23^. Human breast milk contains a diverse lipidome, including phospholipids enriched with long-chain PUFA side chains such as PEP-DHA^11, 24^. ^10^. A recent systematic review found that the breast milk composition was predictive of infant neurodevelopment^25^. Human breast milk is the source of PEP-DHA for young infants, both directly and through precursors (e.g. other ether lipids, EPA)^26^. Notably, ether lipids are abundant in human milk but only in trace amounts in standard cow’s milk-based infant formulas^11^.

Despite strong biological rationale, clinical trials investigating omega-3 fatty acid supplementation and cognition have produced inconsistent findings. Most studies have administered DHA or EPA in esterified rather than ether lipid carriers^27^. A meta-analysis revealed that prenatal maternal DHA/EPA supplementation across these 11 trials showed no improvement in child cognition^28^ and postnatal PUFA supplementation, again without specific consideration of vinyl ether lipid carriers, similarly failed to provide cognitive benefits among bottle-fed and premature infants^29^. These discrepancies suggest that the lipid carrier may influence the bioavailability and biological action of omega-3 fatty acids^8, 24^. It therefore remains to be determined whether PEP-DHA or other ether lipid carriers could enhance neurodevelopmental outcomes relative to conventional esterified forms.

**Extended Data Figure 1.**
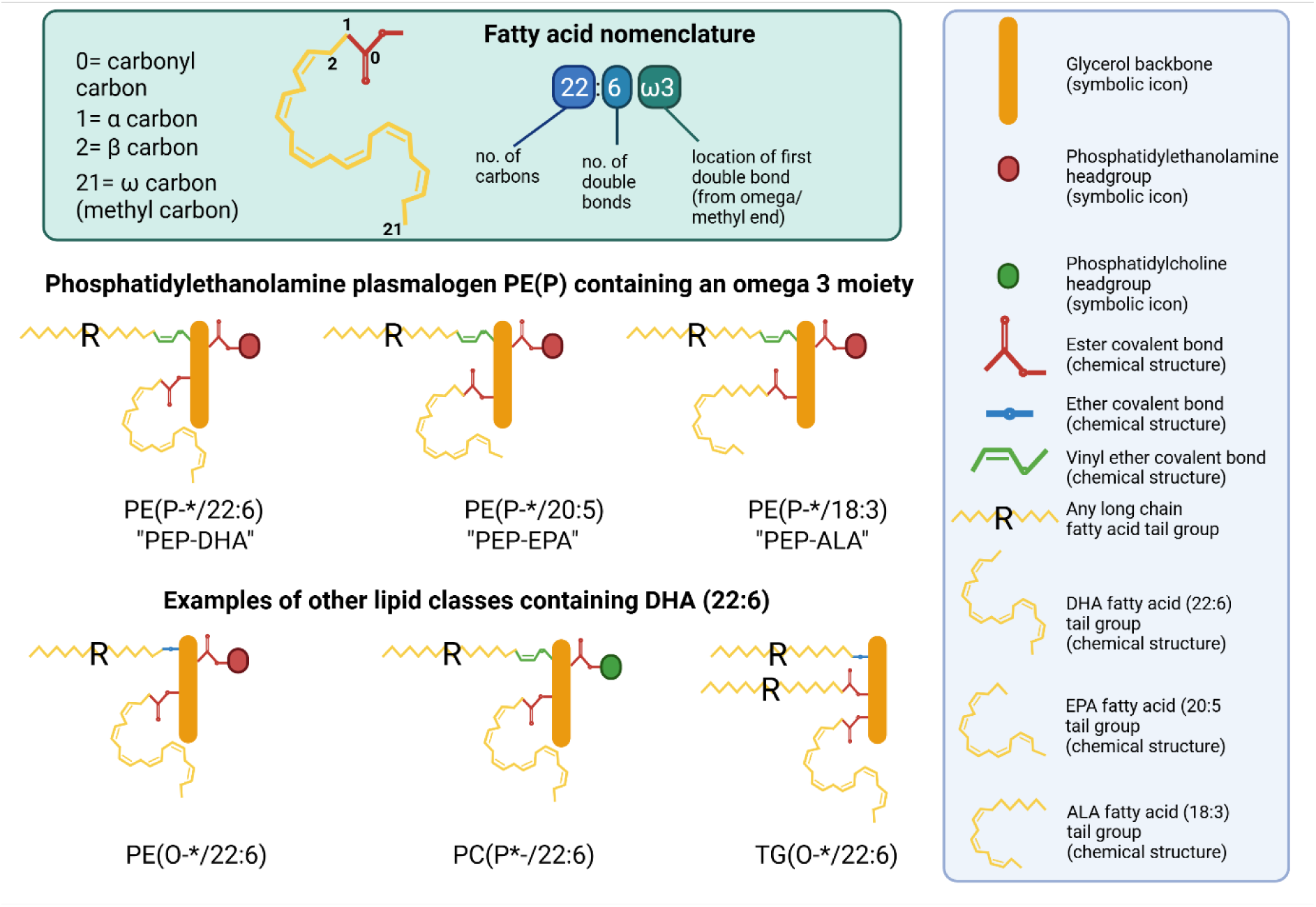
Omega-3–containing plasmalogens and related lipid species. Inset (top left) illustrates fatty-acid notation and carbon numbering from the carbonyl carbon to the ω carbon. Middle row shows omega-3 containing phosphatidylethanolamine plasmalogens, with a vinyl-ether linkage at sn-1 and an omega-3 PUFA at sn-2, left to right PE(P-*/22:6) (PEP-DHA); PE(P-*/20:5) (PEP-EPA); and PE(P-*/18:3) (PEP-ALA). Bottom row depicts related DHA-carrying lipids: plasmanyl-PE (PE(O), “PE(O-*/22:6”; alkyl-ether at sn-1; phosphatidylcholine plasmalogen (PC(P), “PC(P-*/22:6)”, and alkyl-ether triacylglycerol (TG(O), “TG(O*/-22:6)”; one alkyl-ether linkage with two esterified acyl chains) * denotes any acyl or alkyl chain. Made with biorender.com.

Maternal nutrition remains a another key determinant of fetal and infant fatty acid status and the WHO also recommends a high-quality maternal diet during pregnancy^30^ to promote healthy fetal growth and neurodevelopment^31, 32, 33^. High-quality prenatal diet is associated with a range of maternal and infant health benefits^34, 35^. Prenatal maternal diet is a likely driver of infant PEP-DHA levels during pregnancy, with precursors remodelled in tissues as required^12^. The maternal diet is the only source of essential fatty acids for infant development both before and after birth until weaning from breast milk ^5^. Further, postnatal maternal diet will also affect the breast milk fatty acid profile^36^.

The Barwon Infant Study (BIS), an Australian pre-birth cohort of 1074 infants, provides a unique opportunity to examine how maternal diet and infant feeding shape early brain development through metabolomic pathways. We have shown that higher plasmalogen levels are inversely associated with inflammation and infection in infancy, suggesting anti-inflammatory benefits^37^ which relates to neurocognition. We recently reported that poor-quality prenatal diet and maternal inflammation partially mediate the adverse impact of lower socioeconomic position on child cognition and language^35^.

Here, we adopted a life-course approach to investigate lipid-mediated pathways linking maternal nutrition, infant feeding, and early neurodevelopment. Using serial lipidomic measurements from 28 weeks’ gestation to 12 months of age, we: (i) examine how lipid species in maternal and infant serum and plasma are associated with subsequent child Bayley-III cognition and language scores, measured at age 2 years (ii) investigate how maternal diet and breastfeeding contribute to DHA-containing phosphatidylethanolamine plasmalogens (PEP-DHA) levels in prenatal maternal serum and breast milk, and (iii) test whether circulating PEP-DHA levels mediate the beneficial effects of prenatal diet quality and breastfeeding on child cognition and language development.

## Methods

### Study design

BIS is a mother-child longitudinal birth cohort (mothers n=1064; children n=1074; 10 sets of twins) assembled using an unselected antenatal recruitment strategy in the Barwon region of Victoria, Australia^38^. The BIS protocol was approved by the Barwon Health Human Research Ethics Committee (HREC 10/24) and the participating families provided written informed consent.

### Breastfeeding measures

At each interview, mothers were asked if they were breastfeeding and/or formula feeding. If they were not breastfeeding, they were asked to report what date they stopped breastfeeding, from which we calculated the number of weeks the infant was breastfed. Mothers were also asked to report on the date they initiated formula feeding, and the number of weeks their infants were formula fed. Using the date of formula feed initiation, we calculated how many weeks infants were exclusively breastfed and how many weeks were mixed feeding (i.e., mothers were breastfeeding but also supplementing with formula). None of the infants were given any other milk (i.e., cow’s milk) other than human breast milk or infant formula at 6 months or prior. Breastfeeding was categorised into four groups: (1) no breastfeeding, which is exclusive formula feeding (reference category), (2) exclusive breastfeeding (breastfeeding only, no formula), (3) mixed breastfeeding (breastfeeding plus formula), and (4) exclusivity unknown (breastfeeding reported but formula use not specified).

### Prenatal dietary measures

Maternal nutrition over the preceding four weeks was assessed using the Dietary Questionnaire for Epidemiological Studies Version 2 (DQES v2) administered at 28 weeks’ gestation as in our previous studies^39^. The DQES v2 provided data on macronutrients (e.g., fibre), micronutrients (e.g., lutein/zeaxanthin), individual fatty acids, glycaemic index, and fish intake. Derived measures included the Australian Recommended Food Score^40^, the Dietary Inflammatory Index^35, 41^, and principal components (PCs) from a principal component analysis of all food items^42^. As in our previous papers, the PCs reflect broader trends in prenatal maternal diet: PC1, labelled as a ‘modern wholefoods’ dietary pattern, is characterised by higher intakes of nuts, green vegetables, and legumes, alongside lower intakes of white bread, full cream milk, and hamburgers. PC2, labelled as a ‘processed’ dietary pattern, includes higher intakes of pasta, chips, meat, and sweet or salty snacks, and lower intakes of vegetables, tofu, and rye bread^35^. All models including these diet measures, except for the Australian Recommended Food Score (ARFS), were additionally adjusted for total energy intake.

### Serum and plasma lipid measures

Blood lipid measures were obtained from maternal serum samples at 28 weeks’ gestation (n=1032), cord blood serum at birth (n=920), and infant plasma samples at 6 months (n=793) and 12 months (n=735). Maternal and infant blood collection procedures are described elsewhere^26^. Samples were analysed using ultra-high-performance tandem mass spectrometry (UHPLC-MS/MS) lipidomics, and identified 776 lipids from 36 classes as described elsewhere^26^.

### Breast milk lipid measures

Breast milk was collected^11^ at 1 month (n=247) but it was less feasible to collect samples from the 593 mothers still breastfeeding at 6 months (**Table 1**), with samples obtained from 32 participants. The rest for this time point were imputed from 1-month samples (see statistical methods). Mothers were provided with a private room to express milk by hand or breast pump. Pre-feed milk samples were collected from one breast. If mastitis was present in one breast, the opposite breast was used; no breast milk was collected from mothers with mastitis in both breasts; 10–20 mL was collected from each participant and stored in a sterile collection container. Lipidomics was performed utilising ultra-high-performance-tandem mass spectrometry (UHPLC-MS/MS) which identified 979 lipids, as described elsewhere^11^.

**Table 1.**
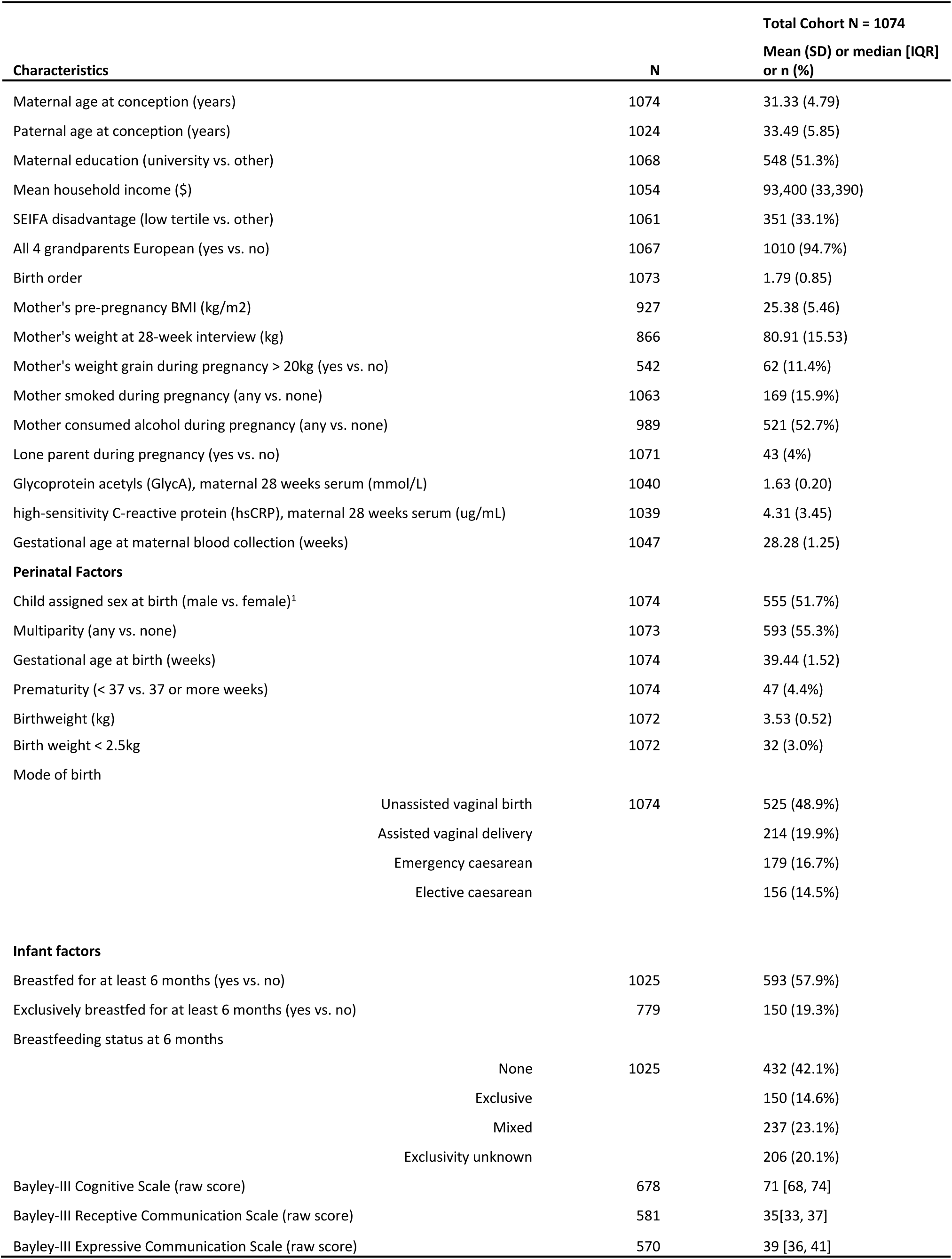
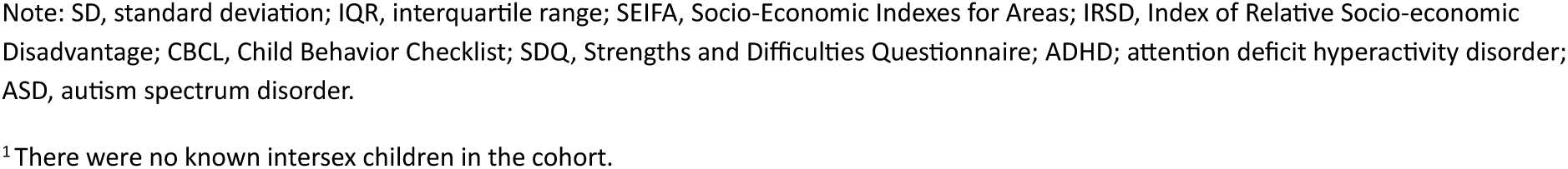
Cohort characteristics.

### Inflammatory measures

Two inflammatory biomarkers, glycoprotein acetyls (GlycA) and high-sensitivity C-reactive protein (hsCRP), were measured and reported previously within the cohort^34, 43^. Maternal non-fasting blood samples were collected at approximately 28 weeks’ gestation. GlycA concentrations (mmol/L) were quantified using metabolomic nuclear magnetic resonance technology (Nightingale Health Ltd., Helsinki, Finland). Levels of hsCRP (mg/L) were determined with a high-sensitivity ELISA assay (Human C-Reactive Protein/hsCRP assay DY1707) as reported in^34, 43^. Zero values were imputed with we added a value equal to 50% of the lowest detected measure for both time points and then the hsCRP variables were log2 transformed^43^.

### Child cognition and language measures

We used the Bayley Scales of Infant and Toddler Development (third edition, Bayley-III). As described elsewhere^34, 44^, a trained research assistant administered the Bayley-III to the children between the ages of 2 and 3 years (mean number of prior assessments completed by RA=139±SD 91.7)^34^. We used the cognition, language, and motor skills subscales. Language was separated into the receptive communication and expressive communication subscales. Motor skills were separated into fine motor and gross motor skills, and analysis involving these subscales are provided in the supplement. Two children were excluded from the analysis: one did not meet the age criteria for the study, and another had significant intellectual disability that was evident prior to the 2-year assessment.

### Statistical approaches

#### Blood lipid weighted gene correlation network analysis (WGCNA)

The dataset was investigated using a data-driven dimensionality reduction approach to identify networks of lipids that were then tested for association with the outcomes. The dimensionality reduction approach we employed was WGCNA, an unsupervised machine-learning method^45^ (**Extended Data Figure 2**), as previously described in our prior work^46^. This systems biology approach, originally devised for uncovering gene networks through co-expression patterns, can identify clusters of lipid concentrations that exhibit co-abundance, potentially indicative of their participation in common biological functions.

Serum and plasma lipid concentrations from 776 lipid features spanning 36 classes were measured using UHPLC-MS/MS over four separate time points – maternal blood at 28 weeks’ gestation, cord blood at birth, infant blood at 6 months, and infant blood at 12 months. These concentrations were log-transformed and standardised into z-scores. A separate WGCNA analysis was conducted on each of the four time points (i.e. data from different timepoints were not pooled). Using the WGCNA package in R (version 4.1.0), a weighted adjacency matrix was generated based on Pearson correlation coefficients of the normalised lipid data. The adjacency matrix was then modified by applying a soft threshold power, selected to strengthen the most significant correlations. The selection of the soft threshold involved plotting the scale-free topology fit index (R²) against a series of potential powers (ranging from 1 to 20), selecting the smallest power at which the index approached a plateau near R² = 0.80, thus adhering to a scale-free network topology. Each soft threshold was specifically tailored to be age-specific to ensure scale-free topology per dataset^45^, with a range from 8 to 10 (maternal serum, 8; umbilical cord serum, 9; infant 6m plasma, 8; infant 12m plasma, 10).

The network’s interconnectedness was assessed by calculating a topological overlap matrix (TOM). Contrary to using solely positive correlations, we chose to analyse unsigned networks to include both positive and negative correlations. This is because in biochemical pathways, negatively correlated metabolites might still interact within the same pathway as either precursors or products. A dendrogram was then constructed using average hierarchical clustering based on a dissimilarity measure (1-TOM), and lipid modules were delineated using a dynamic tree-cutting algorithm. The minimum size for a module was set to 10, with a merging threshold of 0.75 for highly similar modules. The naming convention for WGCNA modules is to assign a number or colour. Each module was associated with an “eigenlipid”, which is the first PC of standardized lipid concentrations in the module, and each sample given a module score by projecting its concentrations onto this PC. The alignment parameter was set to ‘along average’ so that the eigenlipid orientation matched the mean lipid concentration, with positive eigenlipid values corresponding to greater average log lipid concentrations across the module.

#### Lipid network principal component analysis (PCA) scores

Modules formed by WGCNA are unsupervised and data-driven: each lipid is assigned to a single module based on its strongest correlation, and lipid profiles at each timepoint are influenced by different factors. Thus, we performed WGCNA on the lipidomics data at each time point (maternal prenatal, cord blood, 6 months and 12 months) to identify co-expression modules. In addition, for maternal blood we created two extra modules—one for all PEP-DHA species and one for all PEP-EPA species (**Supplementary Table 1**)—by conducting a PCA on each lipid set and taking the first principal component (PC1). In separate minimally adjusted linear regression analyses, we identified which modules were significantly (p<0.05) associated with Bayley-III cognition. The lipid loadings (**Supplementary Dataset 1**) from these cognition-associated modules were then applied to lipid data from the other time points to construct the same modules longitudinally. This allowed us to test whether a given lipid network was related to cognition at one time point but not others, rather than relying on the time-specific eigenlipids generated independently by WGCNA (**Figure 1**). Each module was named by its colour and the time point at which it was originally identified as well as the hub lipid class (e.g., *PC1 MatLightgreen-PEP·DHA(hub)* refers to the Lightgreen module discovered in maternal serum and the hub lipid was PE(P-18:0-22:6), belonging to the class PE(P), *PC1 6MPink-LPC(hub)* refers to the Pink module discovered in the 6 months plasma, and the hub lipid was LPC 18:3, belonging to the class LPC). We also recreated the cognition-associated blood lipid modules in breast milk samples, but instead of using the original loadings, we derived new ones by performing PCA on each corresponding set of breast milk lipids and extracting PC1. As a sensitivity analysis, we repeated this PCA-based approach for the blood lipid modules themselves to evaluate how using new loadings affected the regression results.

**Figure 1.**
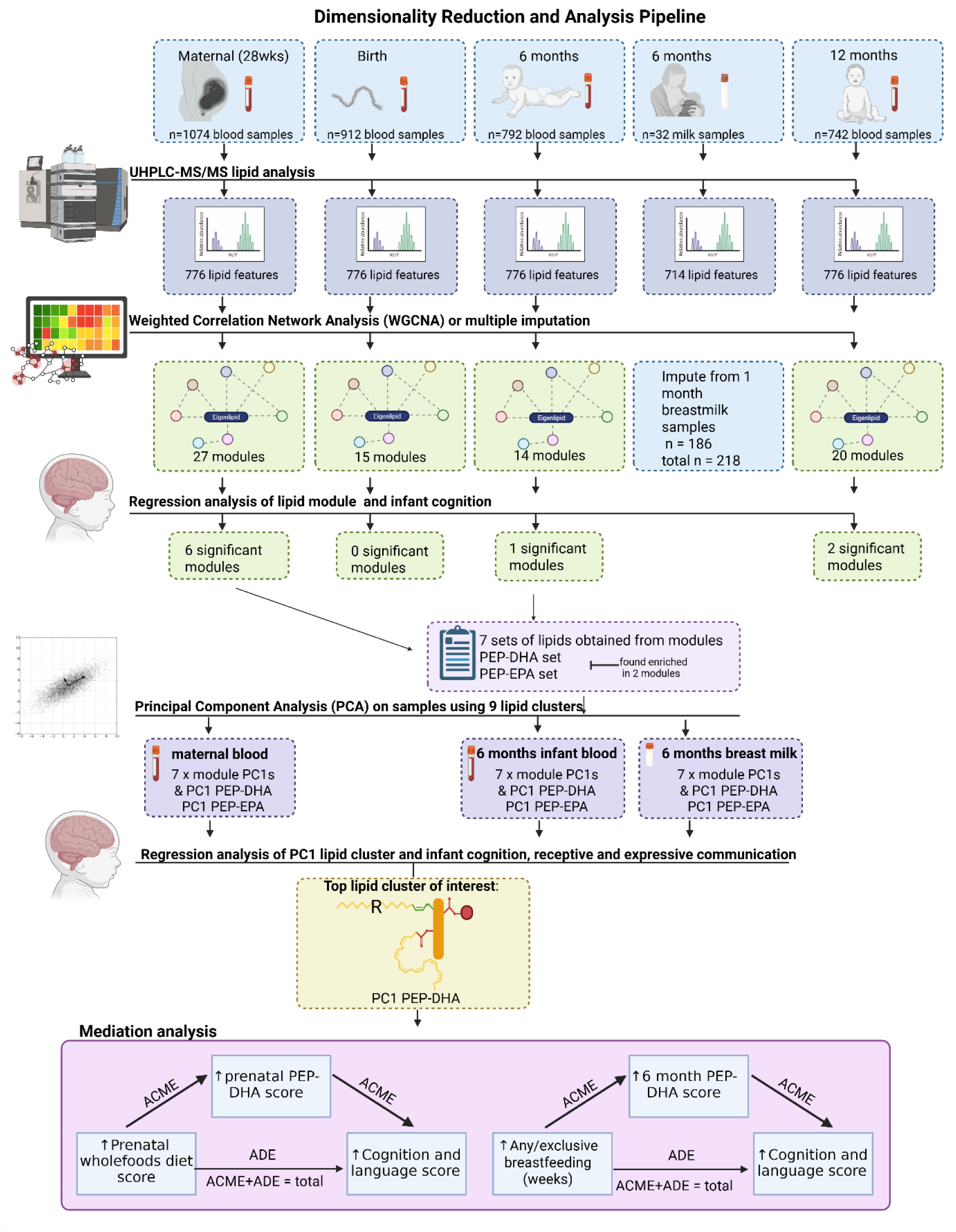
Dimensionality reduction and analysis pipeline. WGCNA was conducted separately on the lipidome from maternal blood at 28 weeks’ gestation as well as infant blood at birth, 6 months and 12 months and regressed against the infant cognition at age 2 years outcome. The lipids that comprised each of the lipid modules that were significantly associated with cognition at the maternal and infant 6 months’ time points were then used to perform Principal Component Analysis (PCA). Applying PCA to these WCGNA derived lipid modules enabled comparison of the same set of lipids across all time points, even if they did not cluster together at every time point. The first principal component (PC1) of PEP-DHA was used in a mediation analysis to determine whether it mediated the relationships between maternal diet or breastfeeding and language and cognition outcomes. Made with biorender.com.

For our main analyses beyond WGCNA, we focused on maternal serum at 28 weeks’ gestation, infant plasma and breast milk at 6 months, as these samples reflect periods where prenatal maternal diet and breastfeeding are key influences. We excluded cord blood because it is strongly affected by delivery and perinatal events^26^ and showed no lipid associations with cognition, and also infant plasma at 12 months, where unmeasured infant diet becomes more influential.

#### Linear regression analyses

Minimally adjusted multivariable linear regression models were used to estimate the strength of association of (i) each of the individual lipid species: maternal (28 weeks’ gestation), infant (6 months old) and breast milk (at 1 month and 6 months), as well as (ii) each of the WGCNA module eigenlipids (in mother at 28 weeks’ gestation, umbilical cord at birth and infant at 6 months and 12 months old) with the Bayley-III subscale outcomes (cognition, receptive communication, and expressive communication). Motor subscales (fine motor and gross motor) results are included in the supplement. Minimal model adjustments included the child’s assigned sex at birth, age at the time of Bayley-III assessment, Bayley-III administrator experience (i.e. number of previous assessments^44^, gestational or child postnatal age at blood collection, minutes from sample collection to storage and sample storage time in days. Fully adjusted models were used to estimate associations on the PCA-derived “Lipid network PCA scores” (maternal (28 weeks’ gestation) and in infant (6 months old)) and the Bayley-III subscale outcomes (**Figure 2**), as well as in the mediation analysis. We also examined the associations between 6-month breast milk (both imputed and non-imputed) lipid network PCA scores and the Bayley-III outcomes (minimally or fully adjusted). The fully adjusted models included the covariates in the minimally adjusted models as well as birth order and maternal prenatal smoking – this was chosen based on knowledge from our past work^34, 35, 46^, and using our data-based approaches^47^. We examined potential confounders for the maternal serum and infant 6-month plasma PEP-DHA and Bayley-III cognition outcome. Potential confounders that did not change the estimate by more than 10% are listed in **Supplementary Table 2**. We also examined whether potential confounders were antecedents, mediators or disease determinants to avoid inappropriate adjustment^47^. Socioeconomic status–related confounders were not included in routine analysis as they are on the causal pathway as an antecedent^34, 35, 48^; however we did conduct a sensitivity analysis including maternal education as a confounder in maternal prenatal and infant (6m) PEP-DHA to Bayley-III cognition associations (**Supplementary Table 3**). Inflammation and diet related variables were explored further as antecedents. Analyses involving the infant’s 6-month blood were additionally adjusted for birthweight. While breastfeeding greatly alters the lipidome at 6 months, we did not adjust for breastfeeding as it is on the causal pathway as an antecedent to infant lipid levels. In each model that was fitted, complete cases were used, except for associations between breast milk lipid PCA scores and the outcomes, which were both complete cases and multiple imputation was used (see imputation methods). Nominal statistical significance was set at p<0.05.

**Figure 2.**
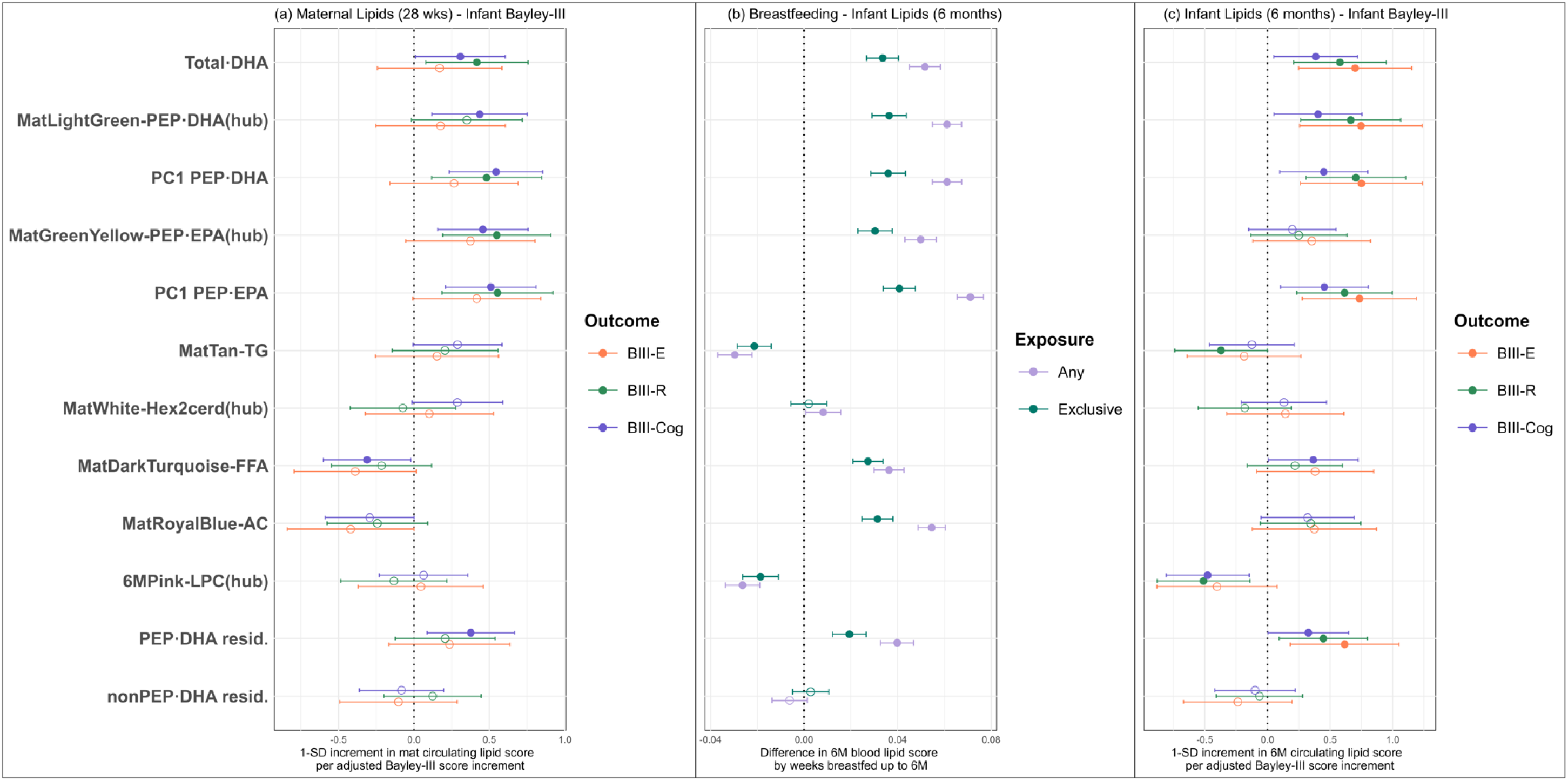
Association between serum lipid network scores in the prenatal maternal (28 weeks’ gestation) measures and in infant plasma at 6 months with child cognition and language at age 2 years, as well as breastfeeding duration. Panel (a) shows the association between each lipid network score in the maternal blood at 28 weeks’ gestation and Bayley-III outcomes. Panel (b) shows association between breastfeeding (any or exclusive) duration by weeks (up to 6 months) and each lipid network score. Panel (c) shows the association between each lipid summary measure in the infant’s blood at 6 months and Bayley-III outcomes. Lipid-Bayley-III models adjusted by child’s assigned sex at birth, age at time of Bayley-III assessment, experience of Bayley-III administrator, (gestational/child) age at blood collection, minutes from blood sample collection to storage, days blood sample stored, birth order, and prenatal maternal smoking. The infant models are additionally adjusted by birthweight. Breastfeeding-Lipid models adjusted by child’s assigned sex at birth, birthweight, child age at blood collection, minutes from blood sample collection to storage, days blood sample stored, birth order, and prenatal maternal smoking. Associations involving the Bayley-III motor scales are found in **Supplementary Figure 3**. M, months; BIII-Cog, Bayley-III cognition scale; BIII-R, Bayley-III receptive communication scale; BIII-E, Bayley-III expressive communication scale; Total DHA, sum of all z-transformed measures species containing DHA, PEP, Phosphatidylethanolamine Plasmalogen; PEP·DHA, PC1 of z-transformed phosphatidylethanolamine plasmalogen containing DHA; PC1 DHA, PC1 of sum of z-transformed lipid species containing DHA. Resid., residual. PEP·DHA resid., the PEP·DHA residual, subtracted from nonPEP·DHA; nonPEP·DHA resid., the nonPEP·DHA residual subtracted from PEP·DHA.

#### Residual analysis

To isolate the specific effects of PEP-DHA independent of the other DHA-containing lipids, we used a residual analysis method. As the PEP-DHA and the sum of all of the other measured species containing a DHA moiety (n=40 species “Total DHA” not including PE(P) species, see list in **Supplementary Table 1**) had partial overlap, we subtracted out the total-nonPEP DHA effect to examine the residual, independent effect for PEP-DHA alone. We first adjusted for total DHA-containing lipids by regressing the sum of the standardised (z-transformed) concentrations of measured PEP-DHA species on the sum of the standardised concentrations of measured non-PEP (“nonPEP DHA”) species containing a DHA moiety. Residuals from this regression represent the variation in PEP-DHA that is not explained by the other DHA-containing lipids. These residuals were then added to the constant term of the regression model to create an adjusted PEP-DHA variable. The list of lipids that comprise these scores can be found in **Supplementary Table 1**.

#### Multiple imputation of 6-month breast milk lipid network PCA scores

A strength of this study was that the breast milk lipidome could also be examined. Breast milk samples used in this study were collected at 1 month (n=186) and 6 months (n=32). Given the small sample of BIS mothers that provided a breast milk sample at 6 months (n=32), we imputed missing values for the 6-month breast milk PEP-DHA score for BIS children who were breastfed at 6 months and whose mothers had provided a breast milk sample at 1 month (n=186). We used multiple imputation by chained equations (mice R package) to generate 90 imputed datasets. The imputation model included variables most strongly correlated with the 6-month breast milk PEP-DHA score (namely, the corresponding 1-month breast milk scores and the maternal prenatal modern wholefoods diet score). In line with multiple imputation guidelines, we also included all variables that were used in subsequent inferential analyses. Regression results from imputed datasets were pooled using Rubin’s rules^49^. For comparison, analysis on complete case data was also conducted.

#### E-value calculations

To assess the robustness of our findings to potential unmeasured confounding, we calculated E-values for the fully adjusted association between maternal PEP-DHA and the odds of being in the lowest quintile of cognition scores vs the other quintiles, as binary measures are required for the E-value calculation^50^. The E-value is the minimum association strength (on the odds-ratio scale) that an unmeasured confounder would need with both the exposure and the outcome—conditional on the measured covariates—to reduce the observed association to the null (OR = 1)^50^. We used the evalue package in Stata to compute the E-value for both the point estimate and the confidence interval^51^.

#### Mediation analysis

We conducted counterfactual mediation analyses to investigate (i) whether associations between prenatal maternal diet quality and child Bayley-III performance at age 2 years were mediated by the maternal PEP-DHA score levels at 28 weeks’ gestation, and (ii) whether associations between breastfeeding at 6 months and child Bayley-III performance at age 2 years were mediated by the PEP-DHA score at 6 months. The plasma lipid network PCA scores (potential mediator) that was associated with breastfeeding (exposure) and with cognition or language (outcome) were each tested as mediators. The total effect consists of two components: the direct effect, which is the relationship between the exposure and outcome not mediated by the lipids, and the indirect effect, which is the exposure-outcome relationship that is mediated by the lipids. Mediation analysis were performed with the *Meflex* package in R. Confidence intervals were obtained using non-parametric bootstrapping, as implemented in medflex.

#### Additional analyses

We previously reported that higher maternal GlycA was linked to lower child Bayley-III cognition and receptive language scores, while better maternal prenatal diet quality was linked to higher scores in these domains^34, 35^. Mediation models were constructed to evaluate whether the maternal PEP-DHA score mediated the relationship between maternal GlycA levels and child Bayley-III cognitive and language scores. As these data are cross-sectional, we also examined the reverse pathway (maternal GlycA as a mediator of PEP-DHA effects). To assess whether prenatal dietary quality exerted effects via PEP-DHA or through inflammation, we included the Australian Recommended Food Score as an exposure variable, examining its associations with maternal GlycA, hsCRP, and PEP-DHA. Mediation analyses tested whether the association between diet quality and outcomes operated through reduced inflammation or enhanced PEP-DHA production.

Longitudinal analyses were performed to estimate maternal and infant PEP-DHA trajectories across pregnancy, 6 months, and 12 months postpartum. To determine whether PEP-DHA exerted independent effects beyond maternal inflammation, associations between maternal PEP-DHA and neurocognitive outcomes were adjusted for diet quality and inflammatory markers (GlycA and hsCRP).

Finally, we compared prenatal PEP-DHA concentrations in mothers who reported vegetarian versus non-vegetarian diets during pregnancy and examined whether maternal vegetarian diet was associated with infant PEP-DHA or neurocognitive outcomes.

## Results

### Cohort characteristics

Participant characteristics are in **Table 1** and participant flowchart in **Supplementary Figure 1**.

Consistent with past literature^19^, any breastfeeding (either combined formula and breast milk feeding or breast milk feeding only) at 6 months was associated with higher scores in the Bayley-III cognition, receptive communication and expressive communication subscales. Each additional week of breastfeeding within the first 6 months was associated with an adjusted mean difference (AMD) of 0.05 higher cognition subscale-adjusted raw score (95% CI 0.02–0.09; *p* = 0.002; **Table 2**). Thus on average, 26 weeks (6 months) of additional breastfeeding corresponds to a 1.3-point higher cognitive scale raw score, equivalent to about a quarter of a standard deviation change.

**Table 2.** The associations between breastfeeding type and duration, breast milk lipids at 6 months, and performance on the Bayley-III at age 2 years.

| Exposure (type of breast milk feeding)<sup>1</sup> | Cognition |  | Receptive communication |  | Expressive communication |  |
| --- | --- | --- | --- | --- | --- | --- |
|  | AMD (95% CI) | p value | AMD (95% CI) | p value | AMD (95% CI) | p value |
| **Continuous measures** |  |  |  |  |  |  |
| Duration (weeks) of breast milk feeding within 6 months | **0.05 (0.02, 0.09)** | **0.002** | **0.04 (0.00, 0.08)** | **0.02** | **0.07 (0.03, 0.12)** | **0.002** |
| Duration (weeks) of exclusive breast milk feeding within 6 months | 0.02 (-0.01, 0.05) | 0.14 | **0.04 (0.00, 0.07)** | **0.04** | 0.03 (-0.01, 0.07) | 0.17 |
| **Categorical measures** |  |  |  |  |  |  |
| Breastfeeding category (ref no breastfeeding at 6 months) |  |  |  |  |  |  |
| Exclusive breast milk feeding for at least 6 months, yes/no | 0.85 (-0.09, 1.79) | 0.07 | **1.31 (0.22, 2.40)** | **0.02** | **1.54 (0.24, 2.83)** | **0.02** |
| Mixed formula and breast milk feeding for at least 6 months, yes/no | **1.05 (0.26, 1.84)** | **0.01** | 0.73 (-0.16, 1.62) | 0.11 | **1.57 (0.50, 2.63)** | **0.004** |
| Breast milk Lipid Score<sup>2</sup> | Cognition |  | Receptive communication |  | Expressive communication |  |
|  | AMD (95% CI) | p value | AMD (95% CI) | p value | AMD (95% CI) | p value |
| PC1 MatLightgreen-PEP·DHA(hub) | 0.3 (-0.03, 0.63) | 0.083 | **0.41 (0.06, 0.76)** | **0.02** | 0.2 (-0.25, 0.66) | 0.39 |
| PC1 PEP·DHA | **0.46 (0.09, 0.84)** | **0.02** | **0.56 (0.17, 0.94)** | **0.007** | 0.22 (-0.3, 0.75) | 0.41 |
| PC1 MatGreenyellow-PEP·EPA(hub) | 0.11 (-0.4, 0.62) | 0.68 | 0.13 (-0.43, 0.69) | 0.65 | -0.14 (-0.88, 0.59) | 0.7 |
| PC1 PEP·EPA | 0.13 (-0.52, 0.78) | 0.7 | 0.19 (-0.52, 0.89) | 0.6 | -0.21 (-1.11, 0.69) | 0.65 |
| PC1 MatWhite-hex2cer(hub) | -0.13 (-0.61, 0.36) | 0.62 | -0.38 (-0.88, 0.12) | 0.14 | -0.31 (-1.03, 0.41) | 0.41 |
| PC1 MatRoyalblue-AC | 0.17 (-0.34, 0.67) | 0.52 | 0.29 (-0.26, 0.85) | 0.31 | 0.2 (-0.47, 0.87) | 0.55 |
| PC1 6MPink-LPC(hub) | -0.02 (-0.4, 0.37) | 0.94 | 0.07 (-0.32, 0.46) | 0.71 | -0.04 (-0.62, 0.53) | 0.89 |

<sup>1</sup>Breastfeeding type and duration models are adjusted by child's assigned sex at birth, age at time of Bayley-III assessment, experience of Bayley-III administrator, birth order, and maternal prenatal smoking.
<sup>2</sup> Breast milk samples (n=32) with additional imputed values (n=186) from mothers who were breastfeeding at 6 months but did not provide a breast milk sample (total n=218). Breast milk lipid-Bayley-III models adjusted by child's assigned sex at birth, child age at breast milk collection, minutes from sample collection to storage, days sample stored.
PC1 – First principal component of a lipid network module identified by WGCNA or by PCA. Module colours (e.g., Lightgreen, Greenyellow, White, Royalblue, Pink) labels represent the different modules. Mat refers to WGCNA conducted in maternal samples, 6M refers to WGCNA conducted in infant (6m) blood. PC1 PEP·DHA, PC1 of all measured phosphatidylethanolamine plasmalogens containing docosahexaenoic acid (22:6n-3). PC1 PEP·EPA, PC1 of all measured phosphatidylethanolamine plasmalogens containing eicosapentaenoic acid (20:5n-3). hex2cer, dihexosylceramide; AC, acylcarnitine; LPC, lysophosphatidylcholine. (hub) indicates the hub lipid species with the highest intramodular connectivity.

### Maternal serum and infant plasma lipid species are individually associated with child Bayley-III cognition scores

We conducted linear regression analyses to estimate the associations between (1) maternal (28 weeks’ gestation) lipids and (2) infant (6 months) lipids and child cognition at age 2 years. In maternal blood, 116/776 (14.9%) lipid species were significantly associated with cognition, with 12 species (all positive associations) maintaining significance at a false discovery rate (FDR) level of 5%. These lipid species were higher levels of Hex2Cer(d16:1/24:1), LPC19:1, TG(50:2 NL-14:0), TG(56:7 NL-20:5), TG(58:8 NL-22:6), Hex2Cer(d16:1/16:0), PE(O)18:1/22:6, PE(O)36:5, PE(P)17:0/22:6, PE(P)18:1/22:6 (A), PE(P)18:1/20:5(A), and PE(P)18:1/20:5(B). Overall, lipid species belonging to classes such as phosphatidylcholine (PC), lysophosphatidylcholine (LPC), phosphatidylethanolamine plasmalogens (plasmenyl-PE (PE(P)), plasmanyl-phosphatidylethanolamine (PE(O)), and dihexosylceramide (Hex2Cer), and some triglycerides (TG) were positively associated with cognition, whereas Ceramide (Cer), Acylcarnitine (AC), and Free Fatty Acid (FFA) were inversely associated. Notably, PE(P) enriched in 22:6 (DHA) and 20:5 (EPA) showed strong positive associations with cognition in maternal blood, with 8 of the top 20 lipid features being PEP-DHA or PEP-EPA (**Supplementary Dataset 3**). In infant blood, 103/776 (13.27%) lipid species were significantly associated with cognition, but none remained significant at an FDR level of 5%. Low levels of PC 18:1/18:1 showed the strongest nominal association (q-value=0.06). (**Supplementary Dataset 4**).

### WGCNA analysis of maternal and infant plasma lipids

Using WGCNA (**Supplementary Dataset 2**), maternal blood was clustered into 29 lipid modules (6 cognition-associated), cord blood into 15 modules (0 cognition-associated), infant 6-month blood into 14 modules (1 cognition-associated) and infant 12-month blood into 20 modules (2 cognition-associated) (**Supplementary Figures 2.1 to 2.4**).

### Maternal prenatal serum lipid network scores at 28 weeks’ gestation are associated with child Bayley-III cognition and language scores

In maternal serum, the two WGCNA lipid modules with PEP hubs (MatLightgreen-PEP·DHA(hub) and MatGreenyellow-PEP·EPA(hub)) were strongly associated with higher infant neurocognition. Key findings include an association between the maternal MatLightgreen-PEP·DHA(hub) module (*p* = 0.007) and the MatGreenyellow-PEP·EPA(hub) module (*p* = 0.002) and higher cognition scores (MatLightgreen-PEP·DHA(hub) AMD = 0.43; 95% CI 0.11–0.75; *p* = 0.007) (**Figure 2**). Given these lipid network modules were found to be enriched in PEP-DHA and PEP-EPA, respectively, we performed a PCA on all measured PEP-DHA species (even those not included in the WGCNA modules MatLightgreen-PEP·DHA(hub) and Greenyellow-PEP·EPA(hub)) to create a PC1 PEP-DHA network score and on PEP-EPA to create a PC1 PEP-EPA network score. The maternal PC1 PEP-DHA and PEP-EPA clusters were positively associated with cognition (AMD = 0.54; *p* = 0.001 and AMD = 0.51; *p* = 0.0009, respectively; **Figures 2 and 3**). Similar patterns were found for receptive communication (**Figure 2**). Lower levels of MatDarkTurquoise-FFA were also associated with infant cognition (AMD = -0.31; 95% CI -0.60–-0.02; *p* = 0.03; **Figure 2**). Higher MatTan-TG, higher MatWhite-Hex2cer and lower MatRoyalBlue-AC were significantly associated with cognition in the minimally adjusted set (**Supplementary Figure 2.1**). These associations were no longer significant after full adjustment, with borderline p values (high MatTan-TG *p* = 0.05; higher matWhite-Hex2cer *p* = 0.061; lower matRoyalBlue-AC *p* = 0.05; **Figure 2**). Lower MatRoyalBlue-AC was also associated with higher Bayley-III gross motor scores (**Supplementary Figure 3**). Higher maternal PEP-DHA was associated with lower adjusted odds of being in the lowest quintile of cognition scores vs the other four quintiles (AOR = 0.77; 95% CI 0.63–0.93; *p* = 0.009). The E-value for the point estimate was 1.91, meaning that an unmeasured confounder would need to have an odds ratio of at least 1.91 with both the exposure and also the outcome to fully explain the observed effect.

**Figure 3.**
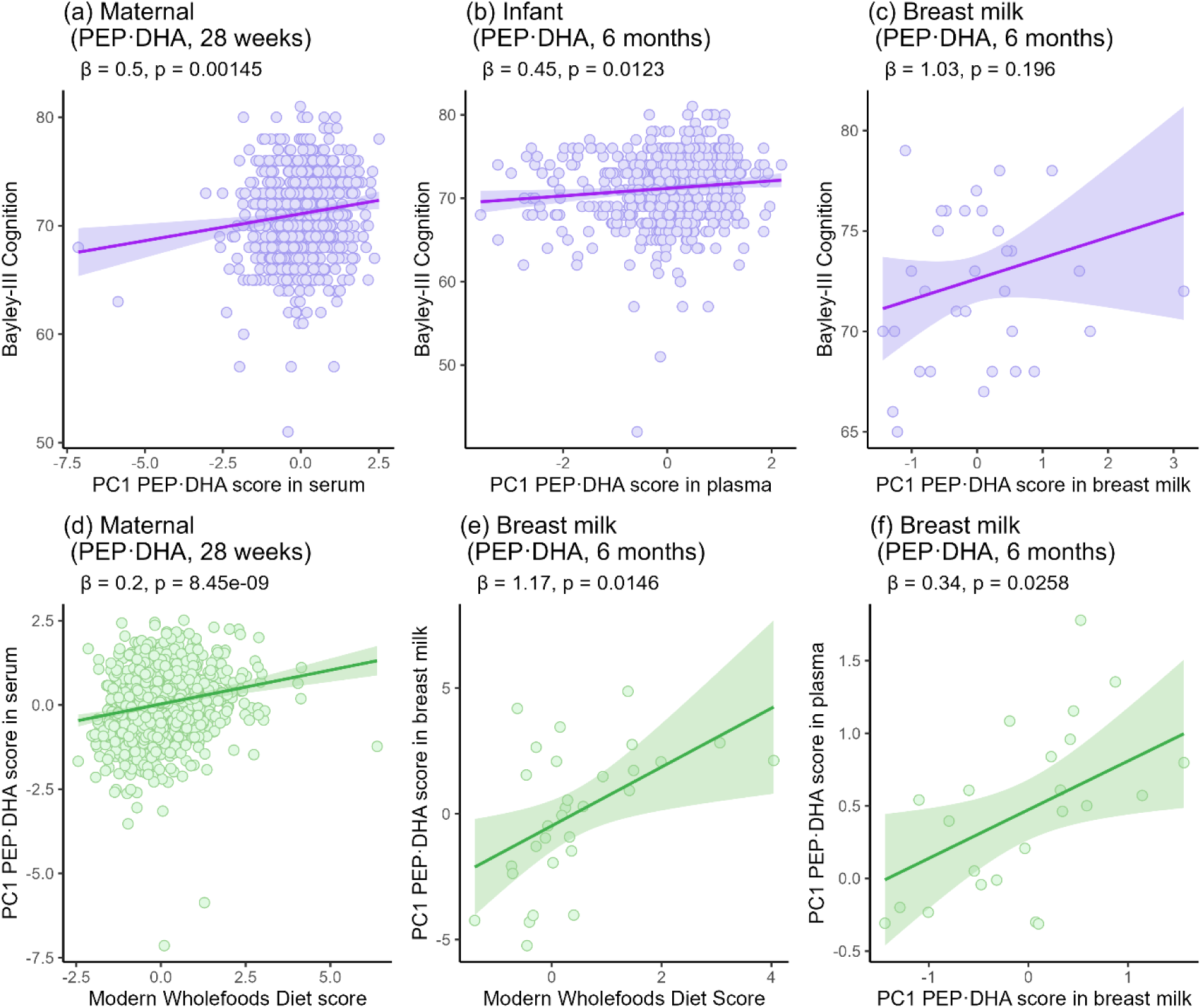
Associations of PEP-DHA lipid scores with child cognition, maternal dietary patterns across maternal, infant, and corresponding lipid measures across maternal, infant, and breast milk samples. The raw cognition score distribution had a mean of 71 and an IQR of 68-74 (Table 1). The top panels represent the association between PEP·DHA lipid scores in **(a)** maternal serum (28 weeks’ gestation), **(b)** infant plasma (age 6 months), and **(c)** breast milk (6 months). Lipid-Bayley-III models adjusted by child’s assigned sex at birth, age at time of Bayley-III assessment, experience of Bayley-III administrator, gestational or child’s age at blood collection, minutes from blood sample collection to storage, days blood sample stored, birth order, and maternal prenatal smoking. The infant model is additionally adjusted by birthweight. The breast milk model is adjusted by child’s assigned sex at birth, child age at breast milk collection, minutes from sample collection to storage, days sample stored. The bottom panels represent the association between prenatal maternal wholefoods diet score and PEP·DHA score in **(d)** maternal serum (28 weeks’ gestation) and **(e)** breast milk (6 months) as well as **(f)** breast milk PEP·DHA and infant plasma PEP·DHA both at 6 months. Modern Wholefoods Diet Score-PEP·DHA models were additionally adjusted for energy intake. Each PEP·DHA measure was z-transformed to aid comparability of effect sizes across models. Each dot represents an individual data point, and shaded areas represent 95% confidence intervals. PC1 PEP·DHA scores use non-imputed data. All associations remained significant after outlier removal using the interquartile range (IQR) method.

### Infant plasma lipid network scores at 6 months are associated with child Bayley-III cognition and language scores

Similar to maternal blood levels, higher PC1 PEP-DHA and PC1 PEP-EPA plasma lipid levels at 6 months were associated with higher scores for all cognition and language outcomes (**Figure 2**). The matLightgreen-PEP·DHA(hub) module and both PC1 PEP-DHA and PC1 PEP-EPA clusters were associated with higher scores for all outcomes (**Figures 2 and 3**). In addition, lower levels of 6MPink-LPC(hub) in the infant 6-month blood were associated with higher cognition and receptive communication scores (AMD = -0.47; *p* = 0.005 and AMD = 0.51, *p* = 0.007, respectively; **Figure 2**). Longer breastfeeding duration (any or exclusive) was associated with higher infant 6-month plasma. PEP-DHA lipid scores (**Figure 2**, middle panel). Across most lipid classes, the direction of association with breastfeeding was similar, indicating a consistent pattern of higher lipid levels among breastfed infants.

Associations between maternal and infant (6 month) lipid network scores and Bayley-III Fine and Gross motor subscales can be found in **Supplementary Figure 3**.

### Health effects are specific to PEP-DHA rather than total PEP plasmalogen or total DHA levels

To isolate the effect of PEP-DHA from that attributable to DHA, we examined the residuals obtained from regressing the standardised sum of PEP-DHA on the standardised sum of non-PEP-DHA and infant cognition. In maternal serum, we found effects of residual PEP-DHA on cognition (p = 0.04), indicating its independent role beyond total DHA levels. At 6 months, the infant residual PEP-DHA was significantly associated with higher cognition (AMD = 0.62; 95% CI 0.18–1.05, *p* = 0.006), receptive language (AMD = 0.45, 95% CI: 0.09–0.80, p = 0.013), and expressive language (AMD = 0.33; 95% CI 0.00–0.65; *p* = 0.047). The reverse, the residuals of a regression of the standardised sum of non-PEP-DHA on standardised sum of PEP-DHA in both maternal and infant blood was not associated with any of the outcomes. Together with the effect size of PEP-DHA being larger than that of Total DHA, it suggests the majority of the effect of circulating DHA is through PEP-DHA (**Figure 2**).

### Infant breast milk lipids are associated with higher child Bayley-III cognition and receptive communication scores

We first examined individual associations between 1-month breast milk and 6-month breast milk lipids (n=714 lipid species) and Bayley-III cognition without imputation, none persisted after FDR correction. At 1 month, the top 3 smallest p-value lipids included TG(26:0), TG(O-52:4) and CE(18:2). 60% of the top 10 lipids were high TG(O) lipids – a type of ether lipid (**Supplementary Dataset 5**). At 6 months, the top 3 most nominally significant lipids were high SM(d18:0/14:0), high FA(22:6) and high FA(18:2) (**Supplementary Dataset 6**). Some 6-month PEP-DHA species showed borderline significance (0.05<*p*<0.10 before FDR at 6 months, but not at 1 month).

Following imputation for missing lipid network score data, a higher level of PC1 of breast milk PEP-DHA at 6 months was associated with higher subsequent cognition (AMD = 0.46; 95% CI 0.09–0.84; *p* = 0.02) as well as receptive communication (**Table 2**, **Figure 3, Supplementary Table 4.1. and 4.2.**). Consistent with this, the WGCNA approach identified the matLightGreen-PEP·DHA (hub) network within breast milk as linked to cognition and receptive communication. We did not find associations between breast milk PEP-DHA (or other networks) at 1 month and any of the Bayley-III outcomes.

### Prenatal maternal dietary patterns are associated with breast milk composition at 6 months

Prenatal diet quality was associated with higher PEP-DHA in the breast milk to at 6 months of infant age. The prenatal maternal modern wholefood dietary pattern was positively associated with higher PEP-DHA levels in breast milk (n=32 at 6 months; AMD = 1.17, 95% CI 0.25–2.08, *p* = 0.01) (**Figure 3**). A similar pattern was found for the Australian Recommended Food Score (AMD = 0.18; 95% CI 0.04–0.33; *p* = 0.02). We also observed similar findings when imputing additional breast milk PEP-DHA data at 6 months (n=186). Further, a higher dietary inflammatory index was associated with lower breast milk PEP-DHA levels at 6 months (AMD = -0.63; 95% CI -0.09-0.36; *p* = <0.0001; **Supplementary Table 5.1. and 5.2.**).

### Maternal levels of circulating PEP-DHA partially mediate the association between maternal dietary patterns and child cognition, receptive communication and expressive communication

We conducted a mediation analysis exploring the effects of maternal dietary patterns during pregnancy on child cognitive and language outcomes; four distinct dietary scores were examined: modern wholefoods dietary pattern, Australian Recommended Food Score, processed dietary pattern, and the Dietary Inflammatory Index (**Table 3 and Supplementary Table 6**). We found partial mediation of these effects through the maternal blood lipid mediator PEP-DHA. There were significant indirect effects of the modern wholefoods diet on cognition (*p _indirect_* = 0.042, **Table 3**), receptive communication (*p _indirect_* = 0.068, **Table 3**), and expressive communication (*p _indirect_* = 0.07, **Table 3**), with the proportions mediated by PEP-DHA around 7% to 10%. This suggests a small but significant influence of prenatal maternal diet through one specific lipid network, PEP-DHA, on child cognitive and language outcomes. Similarly, the Australian Recommended Food Score showed small indirect effects on cognition and communication, with point estimates of the proportion mediated ranging from 9% to 17% **(Supplementary Table 6)**. Conversely, the lower prenatal maternal processed diet scores showed indirect effects on higher cognition, receptive communication, and expressive **(Supplementary Table 6)**. Finally, apparent beneficial effect of lower Dietary Inflammatory Index scores on all outcomes, was also mediated through higher maternal blood PEP-DHA levels **(Supplementary Table 6)**.

**Table 3.**
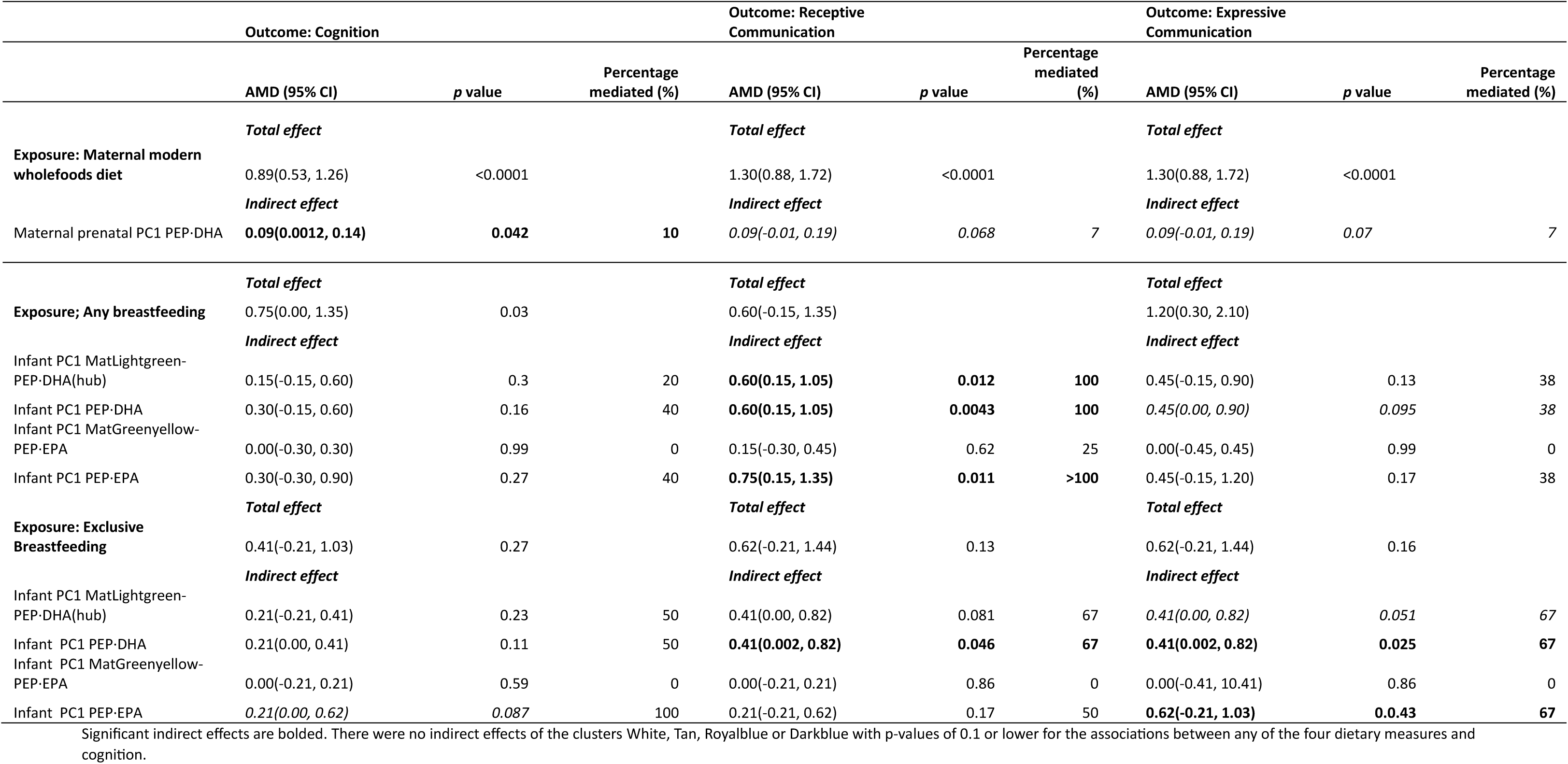
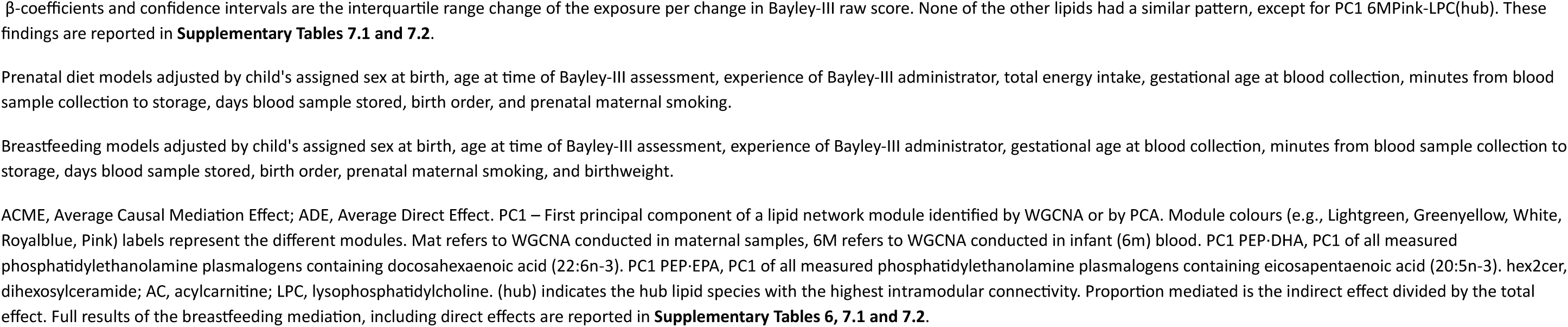
The beneficial effects of prenatal diet quality and breastfeeding are partially mediated through maternal serum and infant plasma (6 months) PEP-DHA and PEP-EPA network lipid levels.

|  | Outcome: Cognition |  |  | Outcome: Receptive Communication |  |  | Outcome: Expressive Communication |  |  |
| --- | --- | --- | --- | --- | --- | --- | --- | --- | --- |
|  | AMD (95% CI) | p value | Percentage mediated (%) | AMD (95% CI) | p value | Percentage mediated (%) | AMD (95% CI) | p value | Percentage mediated (%) |
|  | *Total effect* |  |  | *Total effect* |  |  | *Total effect* |  |  |
| **Exposure: Maternal modern wholefoods diet** | 0.89(0.53, 1.26) | <0.0001 |  | 1.30(0.88, 1.72) | <0.0001 |  | 1.30(0.88, 1.72) | <0.0001 |  |
|  | *Indirect effect* |  |  | *Indirect effect* |  |  | *Indirect effect* |  |  |
| Maternal prenatal PC1 PEP-DHA | **0.09(0.0012, 0.14)** | **0.042** | **10** | 0.09(-0.01, 0.19) | 0.068 | 7 | 0.09(-0.01, 0.19) | 0.07 | 7 |
|  | *Total effect* |  |  | *Total effect* |  |  | *Total effect* |  |  |
| **Exposure; Any breastfeeding** | 0.75(0.00, 1.35) | 0.03 |  | 0.60(-0.15, 1.35) |  |  | 1.20(0.30, 2.10) |  |  |
|  | *Indirect effect* |  |  | *Indirect effect* |  |  | *Indirect effect* |  |  |
| Infant PC1 MatLightgreen-PEP-DHA(hub) | 0.15(-0.15, 0.60) | 0.3 | 20 | **0.60(0.15, 1.05)** | **0.012** | **100** | 0.45(-0.15, 0.90) | 0.13 | 38 |
| Infant PC1 PEP-DHA | 0.30(-0.15, 0.60) | 0.16 | 40 | **0.60(0.15, 1.05)** | **0.0043** | **100** | 0.45(0.00, 0.90) | 0.095 | 38 |
| Infant PC1 MatGreenyellow-PEP-EPA | 0.00(-0.30, 0.30) | 0.99 | 0 | 0.15(-0.30, 0.45) | 0.62 | 25 | 0.00(-0.45, 0.45) | 0.99 | 0 |
| Infant PC1 PEP-EPA | 0.30(-0.30, 0.90) | 0.27 | 40 | **0.75(0.15, 1.35)** | **0.011** | **>100** | 0.45(-0.15, 1.20) | 0.17 | 38 |
|  | *Total effect* |  |  | *Total effect* |  |  | *Total effect* |  |  |
| **Exposure: Exclusive Breastfeeding** | 0.41(-0.21, 1.03) | 0.27 |  | 0.62(-0.21, 1.44) | 0.13 |  | 0.62(-0.21, 1.44) | 0.16 |  |
|  | *Indirect effect* |  |  | *Indirect effect* |  |  | *Indirect effect* |  |  |
| Infant PC1 MatLightgreen-PEP-DHA(hub) | 0.21(-0.21, 0.41) | 0.23 | 50 | 0.41(0.00, 0.82) | 0.081 | 67 | 0.41(0.00, 0.82) | 0.051 | 67 |
| Infant PC1 PEP-DHA | 0.21(0.00, 0.41) | 0.11 | 50 | **0.41(0.002, 0.82)** | **0.046** | **67** | **0.41(0.002, 0.82)** | **0.025** | **67** |
| Infant PC1 MatGreenyellow-PEP-EPA | 0.00(-0.21, 0.21) | 0.59 | 0 | 0.00(-0.21, 0.21) | 0.86 | 0 | 0.00(-0.41, 10.41) | 0.86 | 0 |
| Infant PC1 PEP-EPA | 0.21(0.00, 0.62) | 0.087 | 100 | 0.21(-0.21, 0.62) | 0.17 | 50 | **0.62(-0.21, 1.03)** | **0.043** | **67** |

Significant indirect effects are bolded. There were no indirect effects of the clusters White, Tan, Royalblue or Darkblue with p-values of 0.1 or lower for the associations between any of the four dietary measures and cognition.
$\beta$ -coefficients and confidence intervals are the interquartile range change of the exposure per change in Bayley-III raw score. None of the other lipids had a similar pattern, except for PC1 6MPink-LPC(hub). These findings are reported in **Supplementary Tables 7.1 and 7.2**.
Prenatal diet models adjusted by child's assigned sex at birth, age at time of Bayley-III assessment, experience of Bayley-III administrator, total energy intake, gestational age at blood collection, minutes from blood sample collection to storage, days blood sample stored, birth order, and prenatal maternal smoking.
Breastfeeding models adjusted by child's assigned sex at birth, age at time of Bayley-III assessment, experience of Bayley-III administrator, gestational age at blood collection, minutes from blood sample collection to storage, days blood sample stored, birth order, prenatal maternal smoking, and birthweight.
ACME, Average Causal Mediation Effect; ADE, Average Direct Effect. PC1 – First principal component of a lipid network module identified by WGCNA or by PCA. Module colours (e.g., Lightgreen, Greenyellow, White, Royalblue, Pink) labels represent the different modules. Mat refers to WGCNA conducted in maternal samples, 6M refers to WGCNA conducted in infant (6m) blood. PC1 PEP-DHA, PC1 of all measured phosphatidylethanolamine plasmalogens containing docosahexaenoic acid (22:6n-3). PC1 PEP-EPA, PC1 of all measured phosphatidylethanolamine plasmalogens containing eicosapentaenoic acid (20:5n-3). hex2cer, dihexosylceramide; AC, acylcarnitine; LPC, lysophosphatidylcholine. (hub) indicates the hub lipid species with the highest intramodular connectivity. Proportion mediated is the indirect effect divided by the total effect. Full results of the breastfeeding mediation, including direct effects are reported in **Supplementary Tables 6, 7.1 and 7.2**.

### Circulating levels of PEP-DHA at 6 months mediate the associations between breastfeeding and child cognition and communication at age 2 years

We examined the indirect effects of breastfeeding type (any or exclusive) at 6 months on child cognition, receptive communication, and expressive communication through the mediation of various lipid network scores. Any breastfeeding (duration weeks completed up to 6 months) on receptive communication was mediated through higher PC1 PEP·DHA (proportion mediated=100%, *p _indirect_*=0.006) (**Table 3, Supplementary Table 7.1**). For children exclusively breastfed at 6 months, mediation through PC1 PEP·DHA were notable for receptive communication (*p _indirect_*=0.046) and expressive communication (*p _indirect_*= 0.025), with 67% of the effect mediated in both cases (**Table 3, Supplementary Table 7.2**). These strong findings indicate that PEP·DHA is an important breast milk related lipid for neurocognition. Other lipids also appeared important but to a lesser extent (**Table 3, Supplementary Tables 7.1 and 7.2).**

For any breastfeeding at 6 months, low levels of 6MPink-LPC(hub) partially mediated the effect on cognition (20% mediated, *p* _indirect_= 0.041) and receptive communication (25% mediated, *p_indirect_* = 0.041; **Table 3, Supplementary Table 7.1**). For exclusive breastfeeding at 6 months, 6MPink-LPC(hub) also demonstrated a trend indirect effect on cognition (mediated, *p_indirect_*=0.073), and receptive communication (*p_indirect_*=0.063; **Table 3, Supplementary Table 7.2**), although not significant.

### Additional analysis: (1) Diet, inflammation and PEP-DHA; (2) PEP-DHA is lower in vegetarian mothers

We examined the extent that PEP-DHA may be beneficial due to anti-inflammatory action. We found that the Australian Recommended Food Score acted through higher PEP-DHA to associate with both lower prenatal maternal GlycA and hsCRP, indicating the anti-inflammatory effect of higher prenatal maternal diet quality was partly through higher PEP-DHA levels rather than reduced inflammation. Furthermore, the effect of PEP-DHA on cognition, receptive communication and expressive communication was not solely due to reduced prenatal inflammation because higher maternal PEP-DHA remained associated with all three Bayley-III outcomes after adjusting for maternal GlycA levels.

In BIS, mothers who reported they had a vegetarian diet during pregnancy (6 vegetarian, 1010 non-vegetarian) had lower levels of PEP-DHA in their blood (AMD = -0.81; 95% CI -1.57—-0.32; *p* = 0.04,). The prenatal vegetarian diet showed no significant association with infant PEP-DHA levels or cognitive/language scores. In part this may reflect the low number of women on vegetarian diets during pregnancy.

## Discussion

In this large, population-based prebirth cohort, we identified a consistent association between higher DHA-containing phosphatidylethanolamine plasmalogens (PEP-DHA) levels during pregnancy and infancy and higher cognitive and language outcomes at two years of age. Leveraging a systems biology framework, we combined lipidome-wide screening with mediation analyses to evaluate the causal pathways linking maternal diet, breastfeeding, and neurodevelopment. Elevated PEP-DHA levels in maternal serum and infant plasma mediated a significant portion of the cognitive benefits conferred by high-quality maternal diet and breastfeeding in counterfactual mediation. The consistency of these findings across biospecimens, developmental stages, and neurodevelopmental domains (cognition and language) strengthens the evidence for a biologically meaningful relationship.

These associations were observed across three developmental windows, (pregnancy, six months, and twelve months of age), and across maternal serum, breast milk, and infant plasma samples. Well-established predictors of infant cognition, namely high-quality maternal diet during pregnancy^35^ and breastfeeding^52^, partly mediated their beneficial effects through elevations in circulating PEP-DHA, both prenatally and postnatally. Notably, a dose–response relationship was evident between six-month breast milk PEP-DHA levels and later cognitive and language performance among breastfed infants. Together, these findings suggest that PEP-DHA may represent a mechanistic link through which maternal nutrition and breastfeeding both optimise early neurocognitive development.

The potential benefits of omega-3 fatty acids in early life are frequently emphasized without accounting for the carrier of DHA. Here, PEP-DHA appeared to act independently of total DHA in our study, with residual PEP-DHA levels in maternal serum and infant plasma remaining significantly associated with cognition after adjustment for total (non-PE(P)) DHA. Further, the strength of these association were of higher magnitude. This suggests that the plasmalogen-bound form of DHA, rather than DHA alone, plays a unique and biologically meaningful role in cognitive development.

Previously in this cohort, we demonstrate a higher omega-3 to total fatty acid ratio in maternal serum at 28 weeks gestation was associated with improved cognitive function at two years^34^, yet fish oil supplementation and dietary omega-3 intake did not produce similar effects^34^. Together with the current findings, this may help explain inconsistencies in outcomes of prior omega-3 supplementation trials^28^, most of which utilised non-ether DHA forms as interventions, whereas PEP-DHA may confer distinct neurodevelopmental benefits. Similarly, infant formulas enriched with non-PEP conventional DHA esters show inconsistent findings in enhancement of cognitive outcomes^53, 54, 55^ ^29^. The absence of ether lipids in formula, coupled with higher levels of DHA peroxidation products such as 4-hydroxynonenal, may underlie this limited efficacy for neurocognition compared with breast milk^8^.

Beyond omega-3 plasmalogens, several other lipid networks were associated with child cognition. At 28 weeks’ gestation, four lipid clusters correlated with Bayley-III cognition scores: two positively (Hex2Ceramides and a triglyceride cluster) and two inversely (free fatty acids and acylcarnitines). We and others previously showed that elevated acylcarnitines, linked to inflammation and mitochondrial dysfunction, were also associated with Attention Deficit/Hyperactivity Disorder and Autism Spectrum Disorder symptoms^46, 56^, supporting their role as markers of impaired oxidative metabolism^57, 58^. Postnatally, a lipid network enriched in (lyso)phosphatidylethanolamine, (lyso)phosphatidylcholine, and (lyso)phosphatidylinositol species (18:2, 18:1, and 16:0 moieties) was overrepresented in bottle-fed infants and negatively associated with cognition. This lipid profile further distinguishes breastfed infants and reinforces the neuroactive role for breast milk lipids.

Limitations include limited data on infant diet composition at 6 months within the BIS cohort, though sensitivity analyses indicated that an unmeasured confounder would require an odds ratio ≥1.9 with both prenatal PEP-DHA and child cognition to fully explain the observed association. These findings are also limited to early life neurodevelopment, which is moderately predictive of later executive function^59^, intelligence^60^ and educational attainment^61^. Because six-month breast milk samples were limited, imputation from one-month samples yielded consistent findings. The relatively homogeneous geographic and ethnic composition of BIS may limit generalisability, and replication in more diverse international cohorts is needed. Together with the mediation results, these data support a directional pathway in which dietary intake influences circulating PEP-DHA, which in turn affects neurodevelopment. These findings are compelling but not conclusive. Randomized controlled trials are now required. Dietary plasmalogen-EPA treatment can increase brain DHA content^62^, and recent trials have shown cognitive improvement following plasmalogen supplementation in both animal models and adults with mild Alzheimer’s disease^63^. Given the wide plasmalogen level variability of among breastfeeding mothers and their absence from commercial infant formulas, interventions that elevate PEP-DHA levels, through maternal diet, breastfeeding support, or infant supplementation, should be evaluated for their impact on early neurodevelopment. Such strategies may be particularly valuable for infants from socioeconomically disadvantaged backgrounds with lower maternal diet quality^35^.

In conclusion, DHA-carrying phosphatidylethanolamine plasmalogens (PEP-DHA) have an important role in neurodevelopment and higher antenatal, breast milk and infant postnatal levels were associated with high subsequent cognition and language at age two years. These findings were distinct from total DHA levels alone. Antenatally, maternal PEP-DHA partly mediated the relationship between prenatal diet quality and cognition and language. Postnatally, the beneficial effect of breastfeeding on neurocognition was substantially mediated by PEP-DHA levels in breast milk and also higher PEP-DHA levels in infant plasma at 6 months. These findings suggest that DHA-carrying plasmalogens are key lipid mediators linking early nutrition with optimal neurodevelopment. If confirmed in subsequent human trials, these findings are of high public health significance. The impact of suboptimal maternal diet is likely to be greatest among women with lower socioeconomic status and education, underscoring the need for targeted support in these groups.

## Data availability statement

Barwon Infant Study (BIS) data requests are considered on scientific and ethical grounds by the BIS Steering Committee. If approved, data are provided under collaborative research agreements.

## Acknowledgments

The authors gratefully acknowledge the participants of the Barwon Infant Study (BIS) for their invaluable contributions. We also thank the many current and former cohort staff members.

Thank you to Pranav Adithya, Billi Newton and Tim Chou for assistance with manuscript preparation.

Members of the BIS Investigator Group also include Peter Sly.

## Declaration of Generative AI and AI-assisted technologies in the writing process

During the preparation of this work the first author used ChatGPT-5 for manuscript refinement and copy editing. After using this tool/service, the first author reviewed and edited the content as needed and takes full responsibility for the content of the publication.

