## Supplementary Information for "Higher omega-3 DHA-plasmalogen phospholipid levels across early life mediate the benefits of maternal healthy diet and breastfeeding on infant neurocognition"

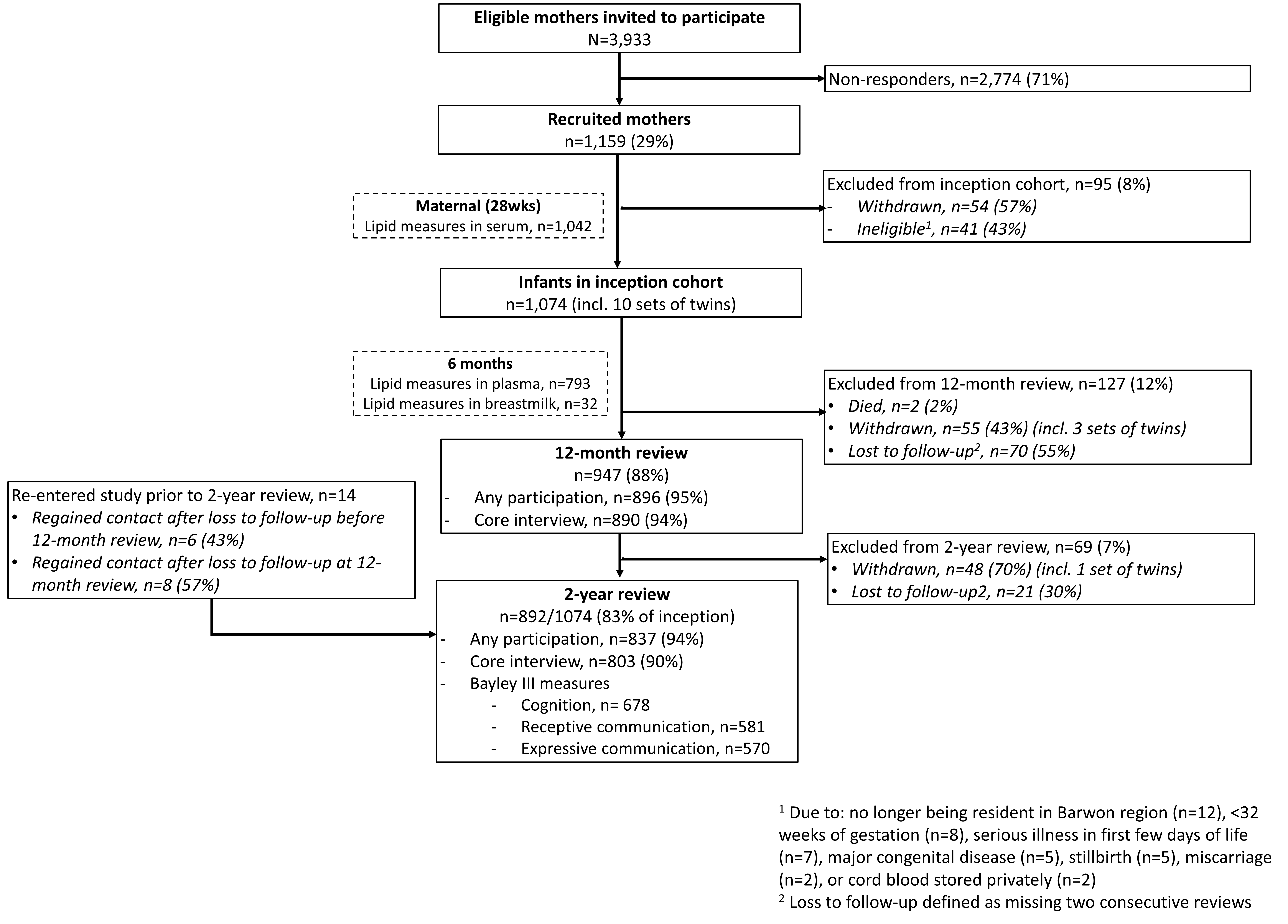

**Supplementary Figure 1. Barwon Infant Study flowchart.** Only participants with complete data for the relevant variables were included in each analysis, unless otherwise noted.

WGCNA Figure variable naming conventions:

Variable names follow the convention z_me[colour]_HS[hubsize]ST[soft-threshold]_timepoint, where z = z-transformed, me[colour] = module eigenlipid (by WGCNA colour), HS[hubsize] = hubsize parameter, ST[soft-threshold] = soft-thresholding power, and timepoint = maternal pregnancy (a), cord blood (CB), 6 months (6M), or 12 months (12M).

See **suplementary dataset 2** for a list of each of the lipid species comprising each module.

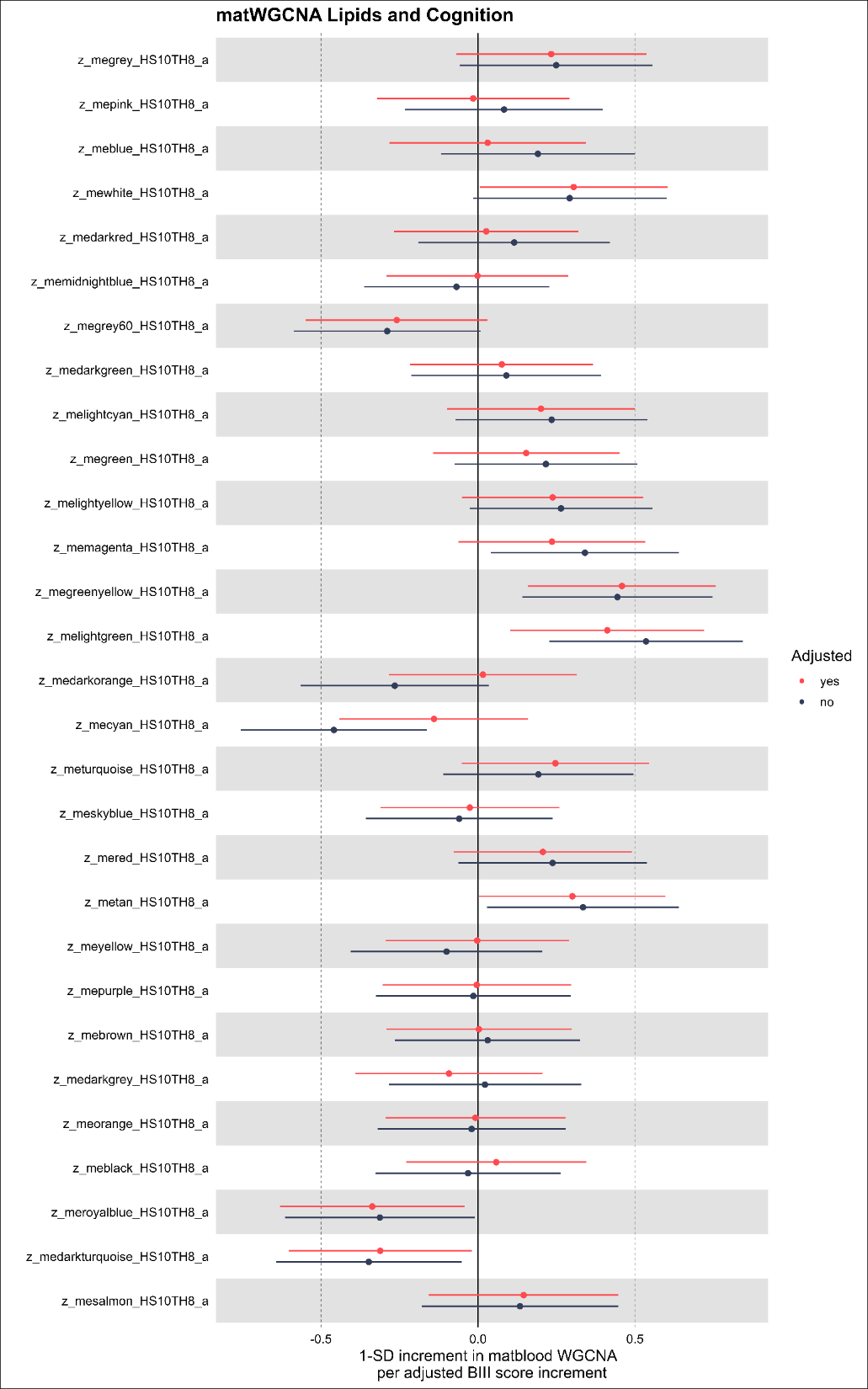

**Supplementary Figure 2.1 Associations between maternal prenatal (28wks) WGCNA modules and Bayley-III cognition.** No-model unadjusted; Yes = models adjusted by child's assigned sex at birth, age at time of Bayley-III assessment, experience of Bayley-III administrator, (gestational/child) age at blood collection, minutes from blood sample collection to storage, days blood sample stored.

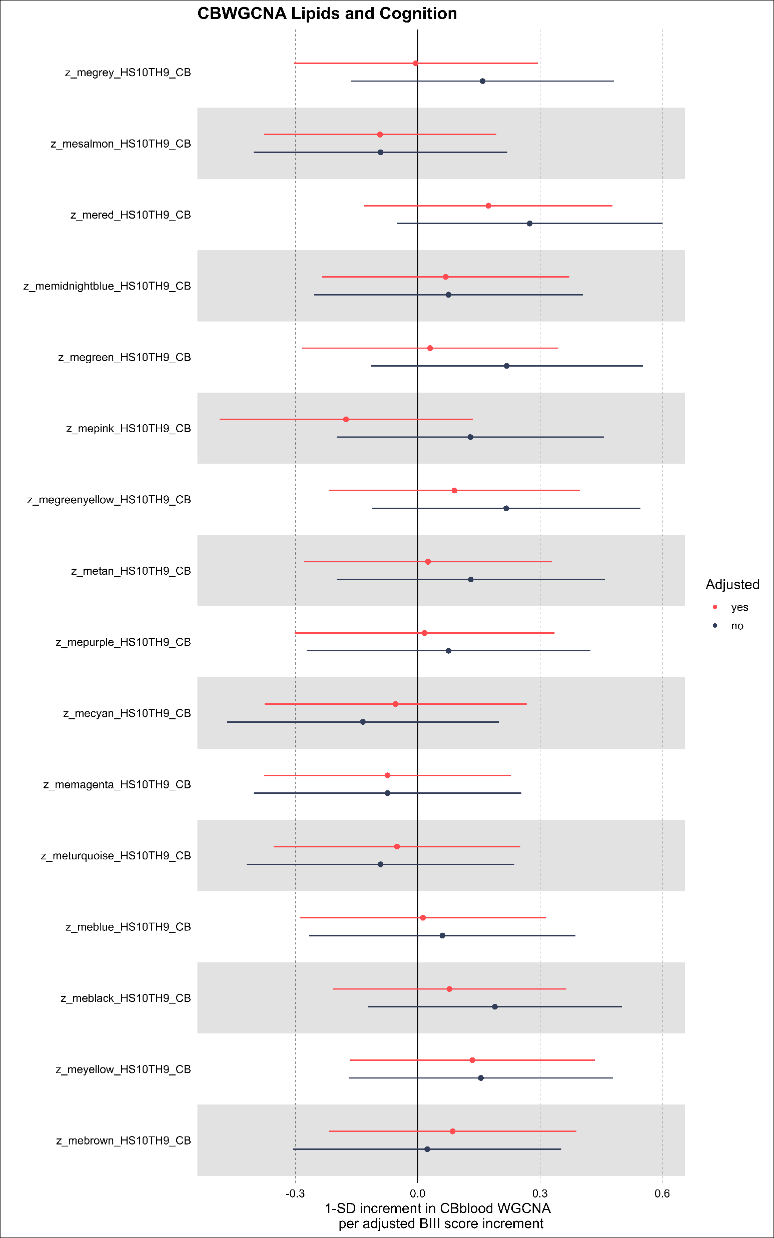

**Supplementary Figure 2.2 Associations between cord blood WGCNA modules and Bayley-III cognition.** No-model unadjusted; Yes = models adjusted by child's assigned sex at birth, age at time of Bayley-III assessment, experience of Bayley-III administrator, (gestational/child) age at blood collection, minutes from blood sample collection to storage, days blood sample stored.
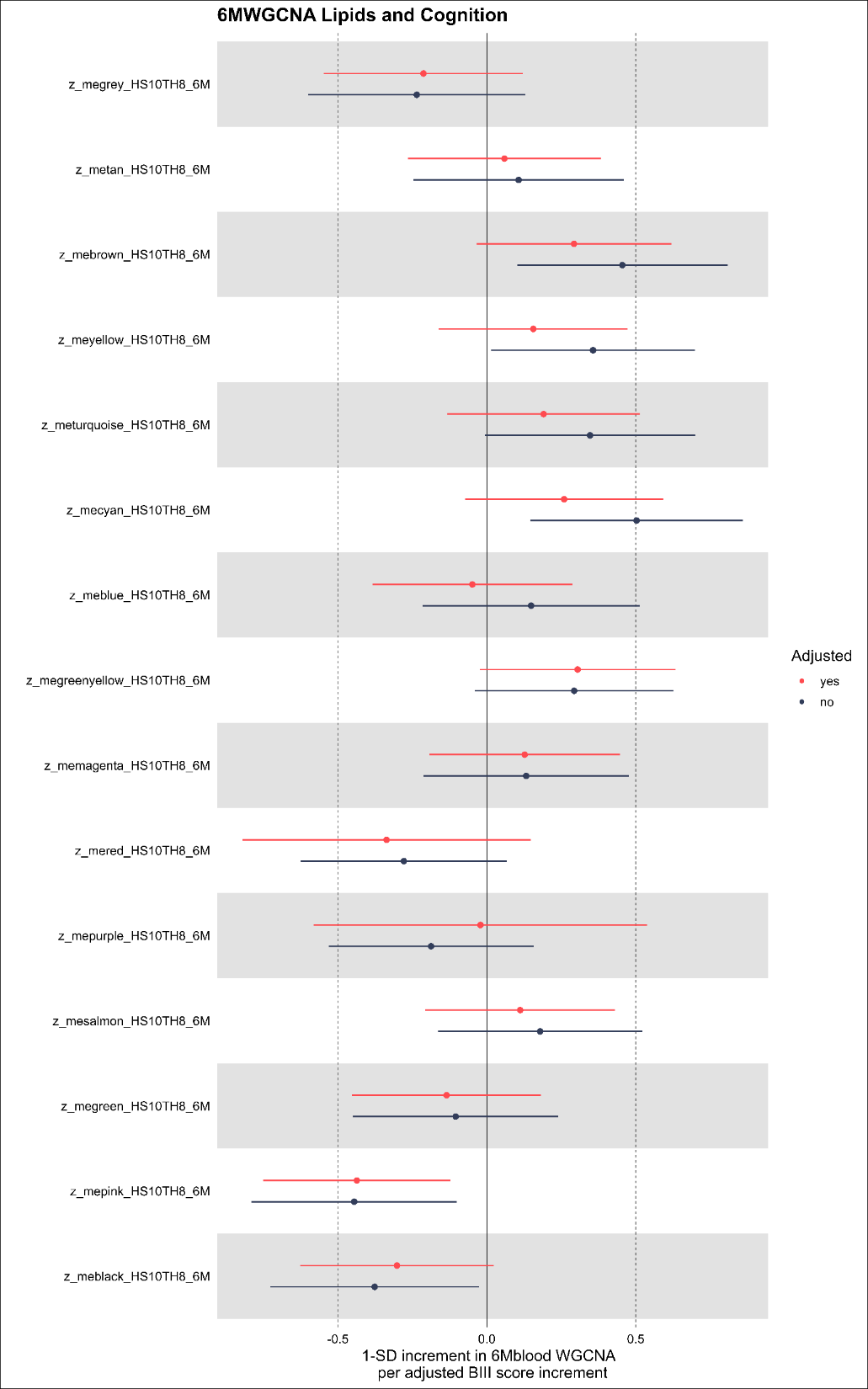

**Supplementary Figure 2.3 Associations between infant (6 months) WGCNA modules and Bayley-III cognition.** No-model unadjusted; Yes = models adjusted by child's assigned sex at birth, age at time of Bayley-III assessment, experience of Bayley-III administrator, (gestational/child) age at blood collection, minutes from blood sample collection to storage, days blood sample stored.

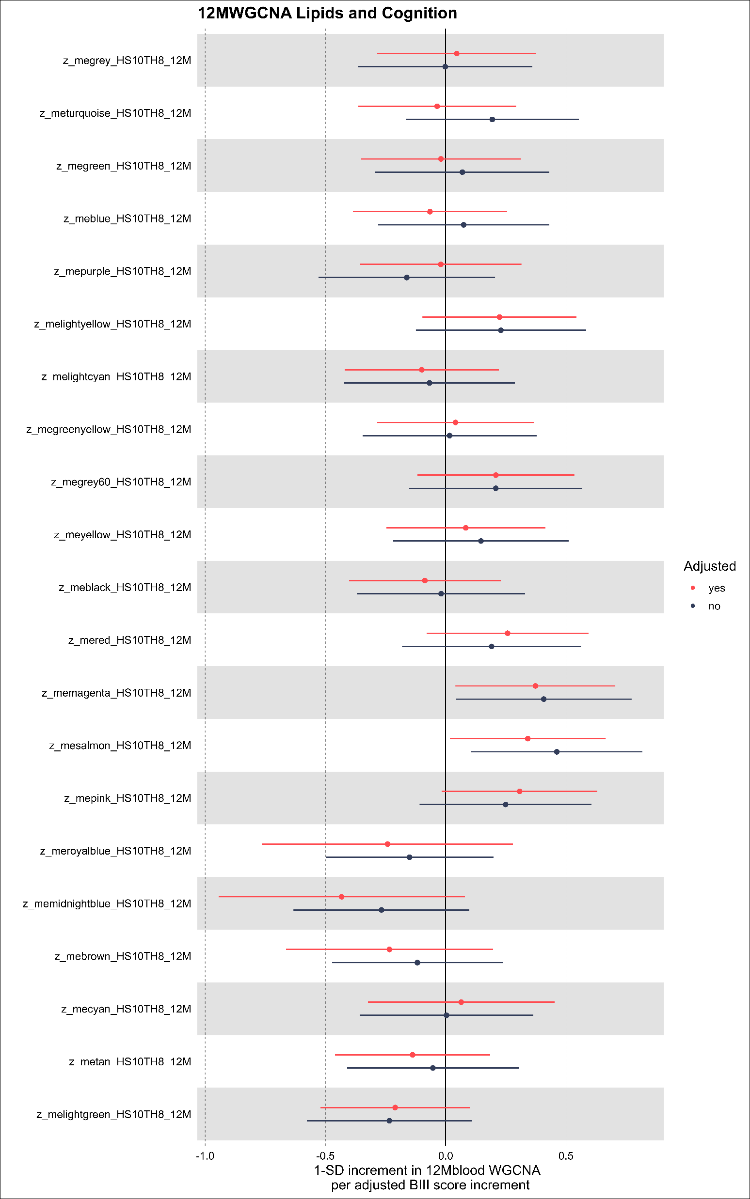

**Supplementary Figure 2.4 Associations between infant (12 months) WGCNA modules and Bayley-III cognition.** No-model unadjusted; Yes = models adjusted by child's assigned sex at birth, age at time of Bayley-III assessment, experience of Bayley-III administrator, (gestational/child) age at blood collection, minutes from blood sample collection to storage, days blood sample stored.

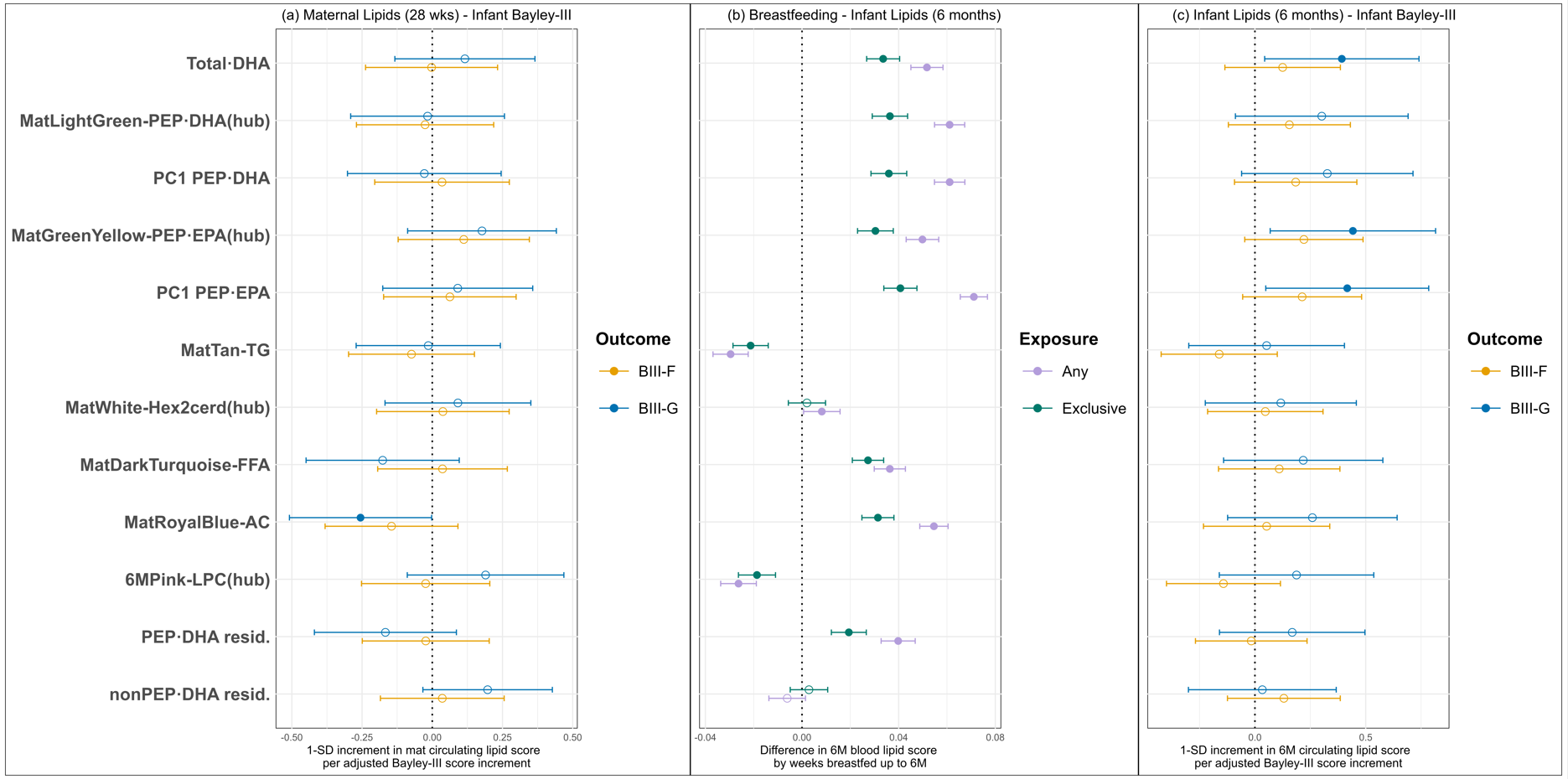

**Supplementary Figure 3. Association between serum lipid summary in the prenatal maternal (28 weeks’ gestation) measures and in infant plasma at 6 months with child Bayley’s-III fine and gross motor scores at age 2 years, as well as breastfeeding duration.**

The left panel shows the association between each lipid summary measure in the maternal blood at 28 weeks’ gestation and Bayley-III outcomes. The middle panel shows association between breastfeeding (any or exclusive) duration by weeks (up to 6 months) and each infant lipid summary measure. The right panel shows the association between each lipid summary measure in the infant’s blood at 6 months and Bayley-III outcomes. Lipid-Bayley-III models adjusted by child's assigned sex at birth, age at time of Bayley-III assessment, experience of Bayley-III administrator, (gestational/child) age at blood collection, minutes from blood sample collection to storage, days blood sample stored, birth order, and prenatal maternal smoking. The infant models are additionally adjusted by child birthweight. Breastfeeding-Lipid models adjusted by child's assigned sex at birth, child birthweight, child age at blood collection, minutes from blood sample collection to storage, days blood sample stored, birth order, and prenatal maternal smoking.

BIII-F, Bayley-III fine motor scale; BIII-G, Bayley-III gross motor scale; Total DHA, sum of all z-transformed measures species containing DHA; PEP, Phosphatidylethanolamine Plasmalogen; PEP·DHA, PC1 of z-transformed phosphatidylethanolamine plasmalogen containing DHA; PC1 DHA, PC1 of sum of z-transformed lipid species containing DHA; resid., residual; PEP·DHA resid., the PEP·DHA residual subtracted from nonPEP·DHA; nonPEP·DHA resid., the nonPEP·DHA residual subtracted from PEP·DHA.
**Supplementary Table 1. List of lipids comprising the PEP-DHA score**

| **PEP-22:6 Score:** | |  |
| --- | --- | --- |
| **The PEP-22:6 score included the following phosphatidylethanolamine plasmalogen species containing 22:6:** | | |
| pep150226a | pep180226 |  |
| pep150226b | pep181226a |  |
| pep160226 | pep181226b |  |
| pep170226a | pep200226 |  |
| pep170226b | pep201226 |  |
| **Total 22:6 Score:** | |  |
| **The total 22:6 score included the following species known to contain 22:6:** | | |
| pc140_226 | peo1802 | dg181_226 |
| pc150_226 | peo1812 | dg182_226 |
| pc160_226 | pep150226a | tg546nl226 |
| pc161_226 | pep150226b | tg547nl226 |
| pc170_226 | pep160226 | tg567nl226 |
| pc180_226 | pep170226a | tg568nl226 |
| pc181_226 | pep170226b | tg569nl226 |
| pco1602 | pep180226 | tg5810nl226 |
| pco1802 | pep181226a | tg588nl226 |
| pcp1602 | pep181226b | tg589nl226 |
| pcp1802 | pep200226 | ce226oh |
| pcp1812 | pep201226 | lpc226oh |
| lpc226sm | lpe226sm |  |
| pe15mho | pi180_226 |  |
| pe160_226 | ce226 |  |
| pe170_226 | de226 |  |
| pe180_226 | fa226 |  |
| pe181_226 | dg160_226 |  |
| peo1602 | dg180_226 |  |

Total DHA -sum of all species included
Total nonPEP DHA - sum of all species, but not including the PEP species in red.

**Supplementary Table 2. Potential confounders that were not included in any models.**

| **These factors did not change the estimate by more than 10%** | |
| --- | --- |
| **Maternal serum prenatal PC1 PEP-DHA score** | **Infant (6months) plasma PC1 PEP-DHA score** |
| Maternal age (years) | Maternal age (years) |
| Paternal age (years) | Paternal age (years) |
| mean household income |  |
| Number of additional household members | Number of additional household members |
| Party more than 1 | Party more than 1 |
| Length at birth | Length at birth |
| Head circumference at birth | Head circumference at birth |

**Supplementary Table 3. Sensitivity analysis including Maternal education (university-educated vs not university educated) as a confounder**

| **Exposure to Bayley-III cognition outcome** | **Covariates** | **AMD (95% CI)** | **p value** |
| --- | --- | --- | --- |
| **Maternal prenatal (28wks)** |  |  |  |
| PC1 PEP-DHA (Prenatal maternal (28wks) serum) | Child's assigned sex at birth, age at time of Bayley-III assessment, experience of Bayley-III administrator, (gestational/child) age at blood collection, minutes from blood sample collection to storage, days blood sample stored, birth order, and prenatal maternal smoking | 0.54 (0.23, 0.85) | 0.001 |
| PC1 PEP-DHA (Prenatal maternal (28wks) serum) | **Maternal education (uni vs no uni)**, child's assigned sex at birth, age at time of Bayley-III assessment, experience of Bayley-III administrator, (gestational/child) age at blood collection, minutes from blood sample collection to storage, days blood sample stored, birth order, and prenatal maternal smoking | 0.49 (0.18, 0.80) | 0.002 |
| **Infant (6 months)** |  |  |  |
| PC1 PEP-DHA (Infant,  6 month plasma) | Child's assigned sex at birth, age at time of Bayley-III assessment, experience of Bayley-III administrator, (gestational/child) age at blood collection, minutes from blood sample collection to storage, days blood sample stored, birth order, and prenatal maternal smoking, child weight at birth | 0.46 (0.10,0.81) | 0.01 |
| PC1 PEP-DHA (Infant,  6 month plasma) | **Maternal education (uni vs no uni)**, child's assigned sex at birth, age at time of Bayley-III assessment, experience of Bayley-III administrator, (gestational/child) age at blood collection, minutes from blood sample collection to storage, days blood sample stored, birth order, and prenatal maternal smoking, child weight at birth | 0.08 (0.01, 0.15) | 0.01 |

**Supplementary Table 4.1. Non imputed.** **The associations between breastfeeding type and duration, breast milk lipids at 6 months, and performance on the Bayley-III at age 2 years**

| **Breast milk** | **Cognition** | |  | **Receptive communication** | |  | **Expressive communication** | |
| --- | --- | --- | --- | --- | --- | --- | --- | --- |
| **Lipid Score** | **AMD (95% CI)** | **p value** |  | **AMD  (95% CI)** | **p value** |  | **AMD  (95% CI)** | **p value** |
| PC1 MatLightgreen-PEP226(hub) | 0.29 (-0.18, 0.76) | 0.21 |  | 0.37 (-0.33, 1.08) | 0.27 |  | 0.24 (-0.53, 1.02) | 0.51 |
| PC1 PEP22:6 | 0.36 (-0.16, 0.88) | 0.16 |  | 0.46 (-0.37, 1.29) | 0.25 |  | 0.21 (-0.66, 1.08) | 0.61 |
| PC1 MatGreenyellow-PEP205(hub) | 0.14 (-0.38, 0.67) | 0.57 |  | 0.36 (-0.49, 1.21) | 0.37 |  | -0.06 (-1.02, 0.89) | 0.88 |
| PC1 PEP20:5 | 0.15 (-0.54, 0.83) | 0.66 |  | 0.38 (-0.72, 1.48) | 0.46 |  | -0.13 (-1.33, 1.08) | 0.82 |
| PC1 MatWhite-hex2cerd(hub) | -0.09 (-0.73, 0.55) | 0.77 |  | -0.36 (-1.23, 0.51) | 0.38 |  | -0.48 (-1.60, 0.64) | 0.37 |
| PC1 MatRoyalblue-AC | 0.16 (-0.47, 0.79) | 0.59 |  | 0.33 (-0.49, 1.15) | 0.39 |  | 0.42 (-0.46, 1.30) | 0.32 |
| PC1 6MPink-LPC(hub) | -0.08 (-0.62, 0.46) | 0.75 |  | 0.45 (-0.28, 1.17) | 0.2 |  | 0.17 (-0.76, 1.10) | 0.7 |
| n=32 samples |  |  |  |  |  |  |  |  |
| **Supplementary Table 4.2. Imputed. The associations between breastfeeding type and duration, breast milk lipids at 6 months, and performance on the Bayley-III at age 2 years** | | | | | | | | |
| **Breast milk** | **Cognition** | |  | **Receptive communication** | |  | **Expressive communication** | |
| **Lipid Score** | **AMD (95% CI)** | **p value** |  | **AMD  (95% CI)** | **p value** |  | **AMD  (95% CI)** | **p value** |
| PC1 MatLightgreen-PEP226(hub) | **0.3 (-0.03, 0.63)** | **0.083** |  | **0.41 (0.06, 0.76)** | **0.029** |  | 0.2 (-0.25, 0.66) | 0.39 |
| PC1 PEP22:6 | **0.46 (0.09, 0.84)** | **0.022** |  | **0.56 (0.17, 0.94)** | **0.0075** |  | 0.22 (-0.3, 0.75) | 0.41 |
| PC1 MatGreenyellow-PEP205(hub) | 0.11 (-0.4, 0.62) | 0.68 |  | 0.13 (-0.43, 0.69) | 0.65 |  | -0.14 (-0.88, 0.59) | 0.7 |
| PC1 PEP20:5 | 0.13 (-0.52, 0.78) | 0.7 |  | 0.19 (-0.52, 0.89) | 0.6 |  | -0.21 (-1.11, 0.69) | 0.65 |
| PC1 MatWhite-hex2cerd(hub) | -0.13 (-0.61, 0.36) | 0.62 |  | -0.38 (-0.88, 0.12) | 0.14 |  | -0.31 (-1.03, 0.41) | 0.41 |
| PC1 MatRoyalblue-AC | 0.17 (-0.34, 0.67) | 0.52 |  | 0.29 (-0.26, 0.85) | 0.31 |  | 0.2 (-0.47, 0.87) | 0.55 |
| PC1 6MPink-LPC(hub) | -0.02 (-0.4, 0.37) | 0.94 |  | 0.07 (-0.32, 0.46) | 0.71 |  | -0.04 (-0.62, 0.53) | 0.89 |
| (n=218 samples. Composed of n=32 original and n=186 imputed) | |  |  |  |  |  |  |  |

**Models adjusted by:** child's assigned sex at birth, child age at breast milk collection, minutes from sample collection to storage, days sample stored.

**Supplementary Table 5.1. Non imputed. The association between maternal prenatal dietary scores and breast milk lipids at 6 months.**

|  | **PC1 PEP-DHA in breast milk (6 months)** | |
| --- | --- | --- |
| **Maternal Dietary Pattern** | **AMD (95% CI)** | **p value** |
| Modern Wholefoods Diet^1^ | **1.17 (0.25, 2.08)** | **0.01** |
| Australian Recommended Food Score | **0.18 (0.04, 0.33)** | **0.02** |
| Processed Dietary Pattern^1^ | -1.30 (-3.70, 1.10) | 0.27 |
| *Inflammatory diet index^1^* | *-0.56 (-1.19, 0.08)* | *0.08* |

n=32 samples

**Supplementary Table 5.2. Non imputed. The association between maternal prenatal dietary scores and breast milk lipids at 6 months.**

|  | **PC1 PEP-22:6 in breast milk (6 months)** | |
| --- | --- | --- |
| **Maternal Dietary Pattern** | **AMD (95% CI)** | **p-value** |
| **Modern Wholefoods Diet^1^** | **1.3 (0.84, 1.76)** | **<0.0001** |
| **Australian Recommended Food Score** | **0.16 (0.09, 0.23)** | **0.00011** |
| Processed Dietary Pattern^1^ | 0.06 (-0.7, 0.82) | 0.88 |
| **Inflammatory Diet Index^1^** | **-0.63 (-0.9, -0.36)** | **<0.0001** |

(n=218 samples. Composed of n=32 original and n=186 imputed)

Models adjusted by: child's assigned sex at birth, age at time of Bayley-III assessment, experience of Bayley-III administrator, total energy intake, gestational age at blood collection, minutes from blood sample collection to storage, days blood sample stored

**Supplementary Table 6. The beneficial effects of prenatal diet quality and cognition/language are partially mediated through PEP-DHA levels.**

| **Modern Wholefoods Diet** | **PEP-22:6** | **Cognition** |  |  | **PEP-22:6** | **Receptive communication** | |  | **PEP-22:6** | **Expressive communication** | |
| --- | --- | --- | --- | --- | --- | --- | --- | --- | --- | --- | --- |
|  | **AMD** | **(95% CI)** | **p value** |  | **AMD** | **(95% CI)** | **p value** |  | **AMD** | **(95% CI)** | **p value** |
| **Direct effect** | 0.65 | (0.35, 0.95) | <0.0001 |  | 0.98 | (0.63, 1.32) | <0.0001 |  | 0.98 | (0.63, 1.32) | <0.0001 |
| **Indirect effect** | **0.07** | **(0, 0.14)** | **0.042** |  | 0.07 | (-0.01, 0.15) | 0.068 |  | 0.07 | (-0.01, 0.15) | 0.07 |
| **Total effect** | 0.72 | (0.43, 1.02) | <0.0001 |  | 1.05 | (0.71, 1.39) | <0.0001 |  | 1.05 | (0.71, 1.39) | <0.0001 |
| **Proportion mediated** | **0.1** |  |  |  | **0.07** |  |  |  | **0.07** |  |  |
| **Australian Recommended Food Score** | **PEP-22:6** | **Cognition** |  |  | **PEP-22:6** | **Receptive communication** | |  | **PEP-22:6** | **Expressive communication** | |
|  | **AMD** | **(95% CI)** | **p value** |  | **AMD** | **(95% CI)** | **p value** |  | **AMD** | **(95% CI)** | **p value** |
| **Direct effect** | 0.05 | (0.01, 0.09) | 0.016 |  | 0.1 | (0.06, 0.14) | <0.0001 |  | 0.1 | (0.06, 0.14) | <0.0001 |
| **Indirect effect** | **0.01** | **(0, 0.01)** | **0.045** |  | **0.01** | **(0, 0.02)** | **0.042** |  | **0.01** | **(0, 0.02)** | **0.042** |
| **Total effect** | 0.05 | (0.02, 0.09) | 0.0059 |  | 0.11 | (0.07, 0.15) | <0.0001 |  | 0.11 | (0.07, 0.15) | <0.0001 |
| **Proportion mediated** | **0.17** |  |  |  | **0.09** |  |  |  | **0.09** |  |  |
| **Processed Dietary Pattern** | **PEP-22:6** | **Cognition** |  |  | **PEP-22:6** | **Receptive communication** | |  | **PEP-22:6** | **Expressive communication** | |
|  | **AMD** | **(95% CI)** | **p value** |  | **AMD** | **(95% CI)** | **p value** |  | **AMD** | **(95% CI)** | **p value** |
| **Direct effect** | -0.75 | (-1.32, -0.18) | 0.0094 |  | -1.18 | (-1.86, -0.5) | 0.00066 |  | -1.18 | (-1.86, -0.5) | 0.00066 |
| **Indirect effect** | **-0.11** | **(-0.22, 0)** | **0.044** |  | **-0.11** | **(-0.22, 0)** | **0.049** |  | **-0.11** | **(-0.22, 0)** | **0.049** |
| **Total effect** | -0.86 | (-1.43, -0.29) | 0.003 |  | -1.29 | (-1.98, -0.61) | 0.0002 |  | -1.29 | (-1.98, -0.61) | 0.00 |
| **Proportion mediated** | **0.13** |  |  |  | **0.09** |  |  |  | **0.09** |  |  |
| **Inflammatory Diet Index** | **PEP-22:6** | **Cognition** |  |  | **PEP-22:6** | **Receptive communication** | |  | **PEP-22:6** | **Expressive communication** | |
|  | **AMD** | **(95% CI)** | **p value** |  | **AMD** | **(95% CI)** | **p value** |  | **AMD** | **(95% CI)** | **p value** |
| **Direct effect** | -0.35 | (-0.54, -0.16) | 0.00027 |  | -0.44 | (-0.65, -0.23) | <0.0001 |  | -0.44 | (-0.65, -0.23) | <0.0001 |
| **Indirect effect** | **-0.04** | **(-0.07, 0)** | **0.04** |  | **-0.04** | **(-0.08, 0)** | **0.03** |  | **-0.04** | **(-0.08, 0)** | **0.03** |
| **Total effect** | -0.39 | (-0.58, -0.2) | <0.0001 |  | -0.48 | (-0.7, -0.27) | <0.0001 |  | -0.48 | (-0.7, -0.27) | <0.0001 |
| **Proportion mediated** | **0.1** |  |  |  | **0.08** |  |  |  | **0.08** |  |  |

**Models adjusted by:** Child's assigned sex at birth, age at time of Bayley-III assessment, experience of Bayley-III administrator, gestational age at blood collection, minutes from blood sample collection to storage, days blood sample stored

PC1 – First principal component of a lipid network module identified by WGCNA or by PCA. Module colours (e.g., Lightgreen, Greenyellow, White, Royalblue, Pink are arbitrary labels from the analysis. Mat refers to WGCNA conducted in maternal samples, 6M refers to WGCNA conducted in infant (6m) blood. PC1 PEP·DHA, PC1 of all measured phosphatidylethanolamine plasmalogens containing docosahexaenoic acid (22:6n-3). PC1 PEP·EPA, PC1 of all measured phosphatidylethanolamine plasmalogens containing eicosapentaenoic acid (20:5n-3). hex2cer, dihexosylceramide; AC, acylcarnitine; LPC, lysophosphatidylcholine. (hub) indicates the hub lipid species with the highest intramodular connectivity.

**Supplementary Table 7.1. The beneficial effects of duration of any breastfeeding and cognition/language are partially mediated through PEP-DHA levels.**

| **PC1 MatLightgreen-PEP226(hub)** | | **Cognition** |  |  |  | **Receptive communication** | |  |  | **Expressive communication** | |
| --- | --- | --- | --- | --- | --- | --- | --- | --- | --- | --- | --- |
|  | **AMD** | **(95% CI)** | **p value** |  | **AMD** | **(95% CI)** | **p value** |  | **AMD** | **(95% CI)** | **p value** |
| **ADE** | 0.04 | (-0.02, 0.09) | 0.19 |  | 0 | (-0.06, 0.06) | 0.95 |  | 0.05 | (-0.02, 0.12) | 0.13 |
| **ACME** | 0.01 | (-0.01, 0.04) | 0.3 |  | **0.04** | **(0.01, 0.07)** | **0.012** |  | 0.03 | (-0.01, 0.06) | 0.13 |
| **Total effect** | 0.05 | (0, 0.09) | 0.03 |  | 0.04 | (-0.01, 0.09) | 0.13 |  | 0.08 | (0.02, 0.14) | 0.0066 |
| **Proportion mediated** | 0.2 |  |  |  | 1 |  |  |  | 0.38 |  |  |
| **PC1 PEP22:6** |  | **Cognition** |  |  |  | **Receptive communication** | |  |  | **Expressive communication** | |
|  | **AMD** | **(95% CI)** | **p value** |  | **AMD** | **(95% CI)** | **p value** |  | **AMD** | **(95% CI)** | **p value** |
| **ADE** | 0.03 | (-0.02, 0.08) | 0.24 |  | 0 | (-0.06, 0.06) | 0.96 |  | 0.05 | (-0.02, 0.12) | 0.13 |
| **ACME** | 0.02 | (-0.01, 0.04) | 0.16 |  | **0.04** | **(0.01, 0.07)** | **0.0043** |  | 0.03 | (0, 0.06) | 0.095 |
| **Total effect** | 0.05 | (0, 0.09) | 0.03 |  | 0.04 | (-0.01, 0.09) | 0.13 |  | 0.08 | (0.02, 0.14) | 0.0066 |
| **Proportion mediated** | 0.4 |  |  |  | 1 |  |  |  | 0.38 |  |  |
| **PC1 MatGreenyellow-PEP205(hub)** | | **Cognition** |  |  |  | **Receptive communication** | |  |  | **Expressive communication** | |
|  | **AMD** | **(95% CI)** | **p value** |  | **AMD** | **(95% CI)** | **p value** |  | **AMD** | **(95% CI)** | **p value** |
| **ADE** | 0.05 | (0, 0.1) | 0.049 |  | 0.03 | (-0.02, 0.09) | 0.22 |  | 0.08 | (0.01, 0.14) | 0.016 |
| **ACME** | 0 | (-0.02, 0.02) | 0.99 |  | 0.01 | (-0.02, 0.03) | 0.62 |  | 0 | (-0.03, 0.03) | 0.99 |
| **Total effect** | 0.05 | (0, 0.09) | 0.03 |  | 0.04 | (-0.01, 0.09) | 0.13 |  | 0.08 | (0.02, 0.14) | 0.0066 |
| **Proportion mediated** | 0 |  |  |  | 0.25 |  |  |  | 0 |  |  |
| **PC1 PEP20:5** |  | **Cognition** |  |  |  | **Receptive communication** | |  |  | **Expressive communication** | |
|  | **AMD** | **(95% CI)** | **p value** |  | **AMD** | **(95% CI)** | **p value** |  | **AMD** | **(95% CI)** | **p value** |
| **ADE** | 0.03 | (-0.03, 0.09) | 0.37 |  | -0.01 | (-0.08, 0.06) | 0.75 |  | 0.05 | (-0.03, 0.13) | 0.25 |
| **ACME** | 0.02 | (-0.02, 0.06) | 0.27 |  | **0.05** | **(0.01, 0.09)** | **0.011** |  | 0.03 | (-0.01, 0.08) | 0.17 |
| **Total effect** | 0.05 | (0, 0.09) | 0.03 |  | 0.04 | (-0.01, 0.09) | 0.13 |  | 0.08 | (0.02, 0.14) | 0.0066 |
| **Proportion mediated** | 0.4 |  |  |  | 1.25 |  |  |  | 0.38 |  |  |
| **PC1 6MPink-LPC(hub)** |  | **Cognition** |  |  |  | **Receptive communication** | |  |  | **Expressive communication** | |
|  | **AMD** | **(95% CI)** | **p value** |  | **AMD** | **(95% CI)** | **p value** |  | **AMD** | **(95% CI)** | **p value** |
| **ADE** | 0.04 | (-0.01, 0.08) | 0.12 |  | 0.03 | (-0.03, 0.08) | 0.32 |  | 0.07 | (0.01, 0.14) | 0.02 |
| **ACME** | 0.01 | (0, 0.02) | 0.041 |  | 0.01 | (0, 0.02) | 0.041 |  | 0.01 | (-0.01, 0.02) | 0.47 |
| **Total effect** | 0.05 | (0, 0.09) | 0.03 |  | 0.04 | (-0.01, 0.09) | 0.13 |  | 0.08 | (0.02, 0.14) | 0.0066 |
| **Proportion mediated** | 0.2 |  |  |  | 0.25 |  |  |  | 0.12 |  |  |

**Models adjusted by:** Child's assigned sex at birth, age at time of Bayley-III assessment, experience of Bayley-III administrator, gestational age at blood collection, minutes from blood sample collection to storage, days blood sample stored

PC1 – First principal component of a lipid network module identified by WGCNA or by PCA. Module colours (e.g., Lightgreen, Greenyellow, White, Royalblue, Pink are arbitrary labels from the analysis. Mat refers to WGCNA conducted in maternal samples, 6M refers to WGCNA conducted in infant (6m) blood. PC1 PEP·DHA, PC1 of all measured phosphatidylethanolamine plasmalogens containing docosahexaenoic acid (22:6n-3). PC1 PEP·EPA, PC1 of all measured phosphatidylethanolamine plasmalogens containing eicosapentaenoic acid (20:5n-3). hex2cer, dihexosylceramide; AC, acylcarnitine; LPC, lysophosphatidylcholine. (hub) indicates the hub lipid species with the highest intramodular connectivity.

**Supplementary Table 7.2. The beneficial effects of duration of exclusive breastfeeding and cognition/language are partially mediated through PEP-DHA levels.**

| **PC1 MatLightgreen-PEP226(hub)** | | **Cognition** |  |  |  | **Receptive communication** | |  |  | **Expressive communication** | |
| --- | --- | --- | --- | --- | --- | --- | --- | --- | --- | --- | --- |
|  |  | **(95% CI)** | **p value** |  | **AMD** | **(95% CI)** | **p value** |  | **AMD** | **(95% CI)** | **p value** |
| **ADE** | 0.01 | (-0.03, 0.05) | 0.6 |  | 0.01 | (-0.03, 0.05) | 0.58 |  | 0.01 | (-0.04, 0.06) | 0.66 |
| **ACME** | 0.01 | (-0.01, 0.02) | 0.23 |  | *0.02* | *(0, 0.04)* | *0.081* |  | *0.02* | *(0, 0.04)* | *0.051* |
| **Total effect** | 0.02 | (-0.01, 0.05) | 0.27 |  | 0.03 | (-0.01, 0.07) | 0.13 |  | 0.03 | (-0.01, 0.07) | 0.16 |
| **Proportion mediated** | 0.5 |  |  |  | 0.67 |  |  |  | 0.67 |  |  |
| **PC1 PEP22:6** |  | **Cognition** |  |  |  | **Receptive communication** | |  |  | **Expressive communication** | |
|  | **AMD** | **(95% CI)** | **p value** |  | **AMD** | **(95% CI)** | **p value** |  | **AMD** | **(95% CI)** | **p value** |
| **ADE** | 0.01 | (-0.03, 0.04) | 0.69 |  | 0.01 | (-0.03, 0.05) | 0.63 |  | 0.01 | (-0.04, 0.06) | 0.68 |
| **ACME** | 0.01 | (0, 0.02) | 0.11 |  | **0.02** | **(0, 0.04)** | **0.034** |  | **0.02** | **(0, 0.04)** | **0.025** |
| **Total effect** | 0.02 | (-0.01, 0.05) | 0.27 |  | 0.03 | (-0.01, 0.07) | 0.13 |  | 0.03 | (-0.01, 0.07) | 0.16 |
| **Proportion mediated** | 0.5 |  |  |  | 0.67 |  |  |  | 0.67 |  |  |
| **PC1 MatGreenyellow-PEP205(hub)** | | **Cognition** |  |  |  | **Receptive communication** | |  |  | **Expressive communication** | |
|  | **AMD** | **(95% CI)** | **p value** |  | **AMD** | **(95% CI)** | **p value** |  | **AMD** | **(95% CI)** | **p value** |
| **ADE** | 0.02 | (-0.02, 0.05) | 0.39 |  | 0.03 | (-0.01, 0.07) | 0.13 |  | 0.03 | (-0.02, 0.08) | 0.2 |
| **ACME** | 0 | (-0.01, 0.01) | 0.59 |  | 0 | (-0.01, 0.01) | 0.86 |  | 0 | (-0.02, 0.02) | 0.86 |
| **Total effect** | 0.02 | (-0.01, 0.05) | 0.27 |  | 0.03 | (-0.01, 0.07) | 0.13 |  | 0.03 | (-0.01, 0.07) | 0.16 |
| **Proportion mediated** | 0 |  |  |  | 0 |  |  |  | 0 |  |  |
| **PC1 PEP20:5** |  | **Cognition** |  |  |  | **Receptive communication** | |  |  | **Expressive communication** | |
|  | **AMD** | **(95% CI)** | **p value** |  | **AMD** | **(95% CI)** | **p value** |  | **AMD** | **(95% CI)** | **p value** |
| **ADE** | 0 | (-0.03, 0.04) | 0.83 |  | 0.01 | (-0.03, 0.05) | 0.47 |  | 0.01 | (-0.04, 0.06) | 0.77 |
| **ACME** | *0.01* | *(0, 0.03)* | *0.087* |  | 0.01 | (-0.01, 0.03) | 0.17 |  | **0.02** | **(0, 0.05)** | **0.043** |
| **Total effect** | 0.02 | (-0.01, 0.05) | 0.27 |  | 0.03 | (-0.01, 0.07) | 0.13 |  | 0.03 | (-0.01, 0.07) | 0.16 |
| **Proportion mediated** | 1 |  |  |  | 0.5 |  |  |  | 0.67 |  |  |
| **PC1 6MPink-LPC(hub)** | | **Cognition** |  |  |  | **Receptive communication** | |  |  | **Expressive communication** | |
|  | **AMD** | **(95% CI)** | **p value** |  | **AMD** | **(95% CI)** | **p value** |  | **AMD** | **(95% CI)** | **p value** |
| **ADE** | 0.01 | (-0.02, 0.04) | 0.52 |  | 0.02 | (-0.02, 0.06) | 0.28 |  | 0.02 | (-0.02, 0.07) | 0.27 |
| **ACME** | *0.01* | *(0, 0.02)* | *0.073* |  | *0.01* | *(0, 0.02)* | *0.063* |  | 0.01 | (0, 0.02) | 0.16 |
| **Total effect** | 0.02 | (-0.01, 0.05) | 0.27 |  | 0.03 | (-0.01, 0.07) | 0.13 |  | 0.03 | (-0.01, 0.07) | 0.16 |
| **Proportion mediated** | 0.5 |  |  |  | 0.33 |  |  |  | 0.33 |  |  |

**Models adjusted by:** Child's assigned sex at birth, age at time of Bayley-III assessment, experience of Bayley-III administrator, gestational age at blood collection, minutes from blood sample collection to storage, days blood sample stored.
