## Supplementary Extended Data Figures for "Higher omega-3 DHA-plasmalogen phospholipid levels across early life mediate the benefits of maternal healthy diet and breastfeeding on infant neurocognition"

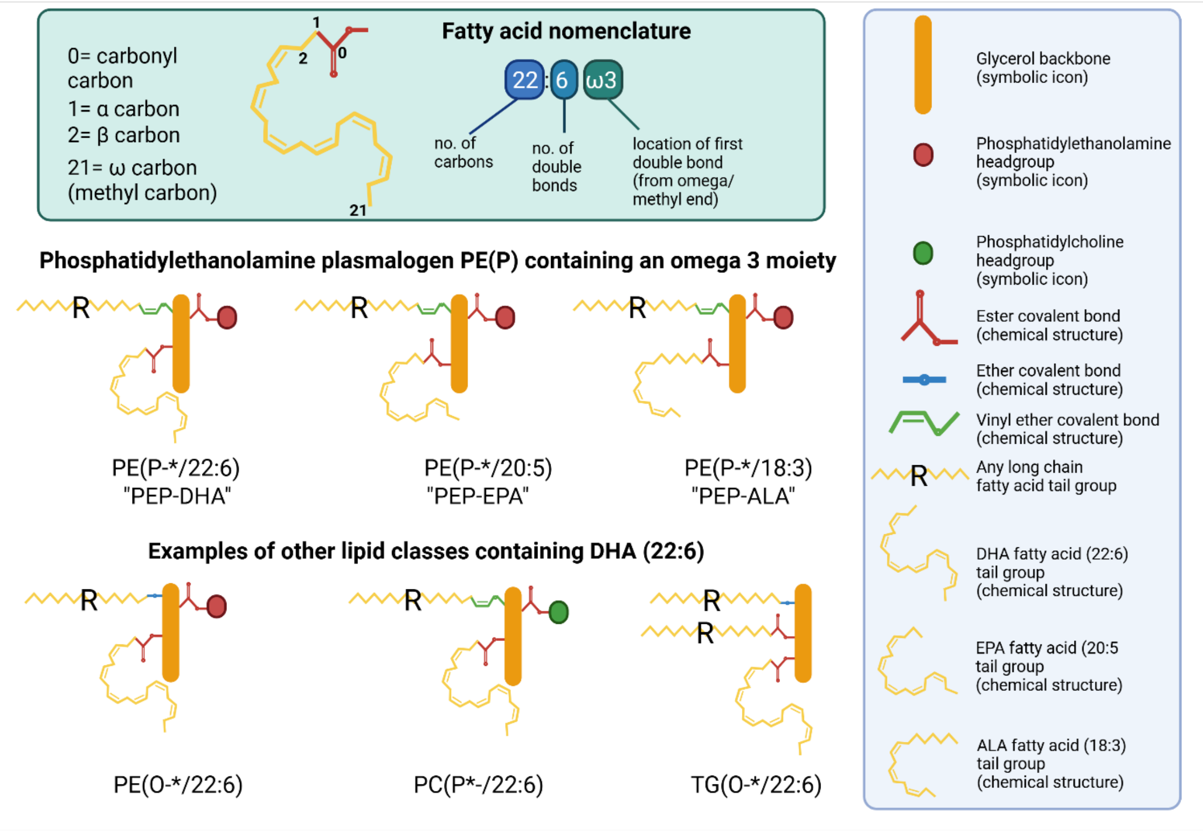


**Extended Data Figure 1. Omega-3–containing plasmalogens and related lipid species.** Inset (top left) illustrates fatty-acid notation and carbon numbering from the carbonyl carbon to the ω carbon. Middle row shows omega-3 containing phosphatidylethanolamine plasmalogens, with a vinyl-ether linkage at sn-1 and an omega-3 PUFA at sn-2, left to right PE(P-*/22:6) (PEP-DHA); PE(P-*/20:5) (PEP-EPA); and PE(P-*/18:3) (PEP-ALA). Bottom row depicts related DHA-carrying lipids: plasmenyl-PE (PE(O), "PE(O-*/22:6"; alkyl-ether at sn-1; phosphatidylcholine plasmalogen (PC(P), "PC(P-*/22:6)", and alkyl-ether triacylglycerol (TG(O), "TG(O*/-22:6)"; one alkyl-ether linkage with two esterified acyl chains) * denotes any acyl or alkyl chain.

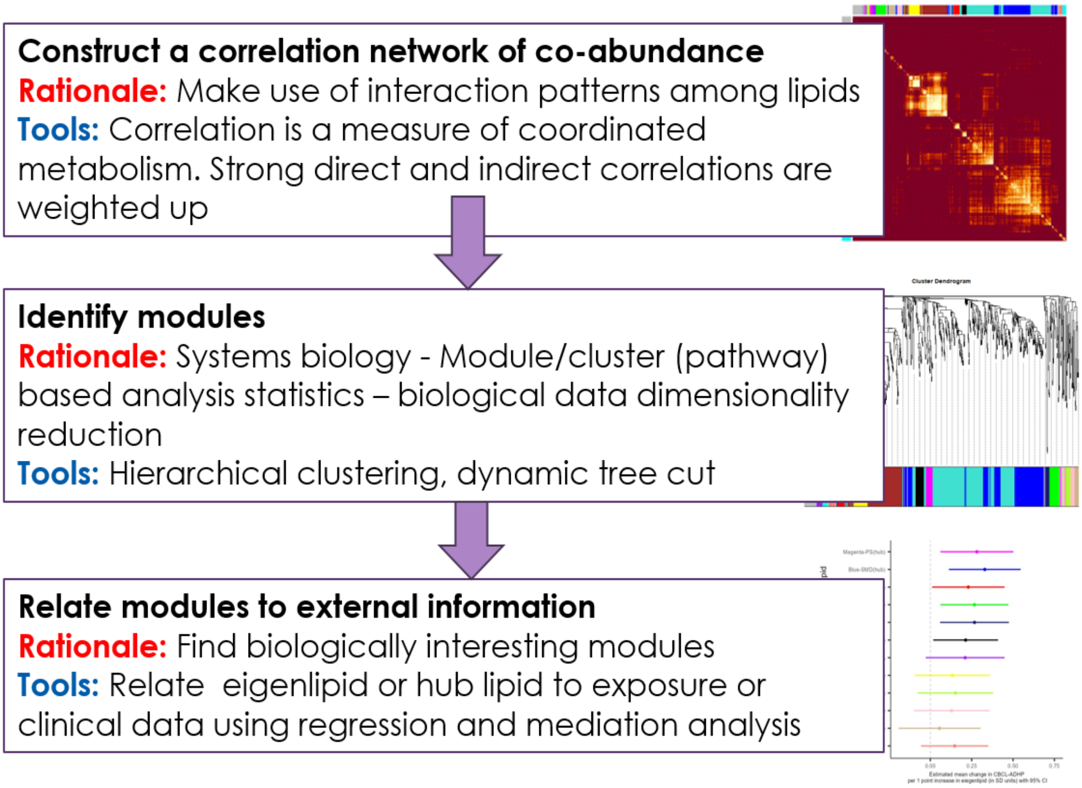


**Extended Data Figure 2.** WGCNA for lipidomic data. An overview of the main steps of weighted gene correlation network analysis for lipid concentration data. Example images from Vacy et al. (2024). Diagram adapted from Langfelder and Hovarth (2008)
